# Phenome-wide Investigation of Behavioral, Environmental, and Neural Associations with Cross-Disorder Genetic Liability in Youth of European Ancestry

**DOI:** 10.1101/2023.02.10.23285783

**Authors:** Sarah E. Paul, Sarah M.C. Colbert, Aaron J. Gorelik, Isabella S. Hansen, I. Nagella, L. Blaydon, A. Hornstein, Emma C. Johnson, Alexander S. Hatoum, David A.A. Baranger, Nourhan M. Elsayed, Deanna M. Barch, Ryan Bogdan, Nicole R. Karcher

## Abstract

Etiologic insights into psychopathology may be gained by using hypothesis-free methods to identify associations between genetic risk for broad psychopathology and phenotypes measured during adolescence, including both markers of child psychopathology and intermediate phenotypes such as neural structure that may link genetic risk with outcomes. We conducted a phenome-wide association study (phenotype n=1,269-1,694) of polygenic risk scores (PRS) for broad spectrum psychopathology (i.e., Compulsive, Psychotic, Neurodevelopmental, and Internalizing) in youth of PCA-selected European ancestry (n=5,556; ages 9-13) who completed the baseline and/or two-year follow-up of the ongoing Adolescent Brain Cognitive DevelopmentSM (ABCD) Study. We found that Neurodevelopmental and Internalizing PRS were significantly associated with a host of proximal as well as distal phenotypes (Neurodevelopmental: 187 and 211; Internalizing: 122 and 173 phenotypes at baseline and two-year follow-up, respectively), whereas Compulsive and Psychotic PRS showed zero and one significant associations, respectively, after Bonferroni correction. Neurodevelopmental PRS were further associated with brain structure metrics (e.g., total volume, mean right hemisphere cortical thickness), with only cortical volume indirectly linking Neurodevelopmental PRS to grades in school. Genetic variation influencing risk to psychopathology manifests broadly as behaviors, psychopathology symptoms, and related risk factors in middle childhood and early adolescence.

## MAIN

Psychopathology is common, highly comorbid, heritable, and associated with a host of negative outcomes (e.g., increased morbidity and mortality)^1–3^. The high polygenicity of and genetic covariance among psychiatric disorders parallel the large degree of symptom and diagnostic overlap at the phenotypic level ^4^. Although progress has been made in characterizing the patterns of comorbidity and consolidating new and existing findings into shared frameworks of overarching functional domains (e.g., HiTOP,^5^ RDoC^6^), etiological and mechanistic insight needed for prevention and mitigation efforts is still limited.

Recent advances in genomic methodologies follow decades of clinical phenotype refinement, shifting the focus from discrete diagnostic categories to higher order symptom clusters. With recently developed techniques, such as Genomic Structural Equation Modeling (Genomic SEM)^7^, summary statistics from Genome-wide Association Studies (GWAS) of psychiatric phenotypes can be leveraged to identify genomic variants associated with higher order dimensions of psychopathology. Both phenotypically and genetically, transdiagnostic factors offer improved validity and power to identify genomic risk loci and associations with other traits of interest, particularly as GWAS sample sizes increase and further advance the utility of Genomic SEM ^7,8^. Polygenic scores (PRS) generated from multivariate GWAS tend to explain more variance than those from univariate GWAS, enhancing their value as proxies for overall genetic liability ^7^. Better power and predictive utility of PRS enhances the likelihood of identifying potential intermediate phenotypes observed in childhood and adolescence that may link genetic susceptibility to psychopathology outcomes, thus improving mechanistic insight. For instance, studies have begun to investigate neural metrics as possible mechanisms driving associations between genetic liability and phenotypic expression ^e.g., 9,10^.

Despite novel insights gained by hypothesis-driven psychopathology research with PRS (e.g., that ADHD PRS is associated with increased screen time in youth^11^), such work is constrained in its potential scope. Further etiological insights can be gained using hypothesis-free methods, such as phenome-wide association studies (PheWAS), which explore associations between genetic risk and hundreds or thousands of phenotypes spanning multiple domains^12^. Effects that survive correction for multiple testing may replicate previously discovered genotype-phenotype associations or identify novel associations that may further drive the development of new hypotheses and theory.

Here, we use Genomic SEM to replicate a recent model of cross-disorder genetic risk and conduct a multivariate GWAS using the largest available GWAS summary statistics of 11 psychiatric disorders. We then compute PRS of the resulting latent genetic factors in the PCA-selected European ancestry subsample of the independent Adolescent Brain Cognitive Development (ABCD) Study sample, from which we conduct a PheWAS testing associations between each PRS with phenotypes measured at the baseline (sample n = 5,556; phenotype n = 1,269; ages 9-10) and two-year-follow-up (sample n = 5,048; phenotype n = 1,694; ages 10-13) assessment waves. We conceptualized these phenotypes as being of multiple types: a) more <u>proximal</u> phenotypes that reflect symptoms of various disorders as well as potentially trait-like characteristics that might also reflect psychopathology or related risk factors (e.g., behavioral inhibition); and b) more <u>distal</u> phenotypes that might reflect behavior related to psychopathology or correlated features (e.g., electronic media use, physical health, cognition, etc., **Supplemental Tables S1-S4**). To further mechanistic insight, we also explore whether baseline neural metrics indirectly link PRS to follow-up outcomes (**Supplemental Table S5)**. The young age of this deeply phenotyped sample allows us to investigate early associations with subthreshold symptoms and related traits largely preceding the development of functional impairment, providing etiological insights beyond what is achievable with Electronic Health Records that may prove fruitful for clinical applications that involve early intervention or even prevention.

## RESULTS

See **Figure 1** for a summary of the analytic process.

**Figure 1.**
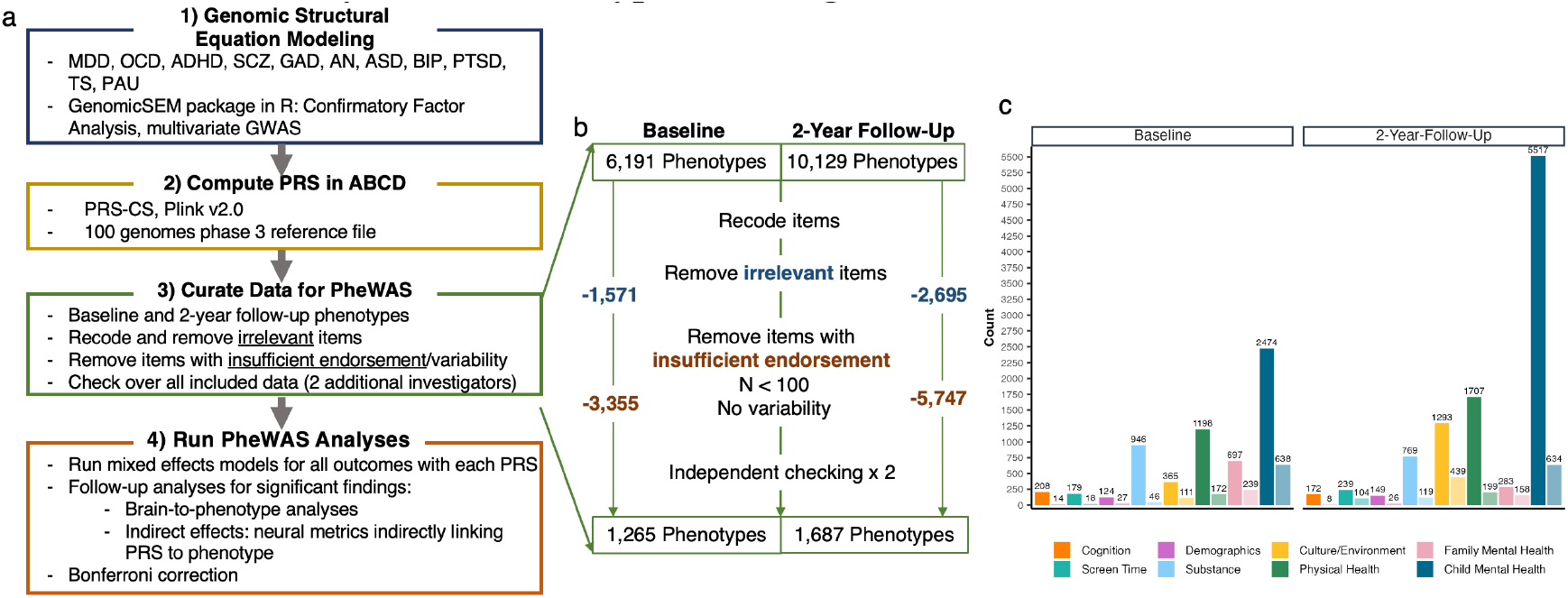
Data Analysis and Phenotype Curating Flow Chart. **a**. Flow chart depicting data analytic process. **b**. Flow chart depicting curating of phenotypes in the Adolescent Brain Cognitive Development (ABCD) dataset. **c**. Bar plot illustrating the total number of phenotypes per domain considered for analysis and retained for the final PheWAS. The left and right panels depict phenotypes at baseline and two-year follow-up assessment waves, respectively. The y-axis reflects the number of phenotypes, and the x-axis shows the domains of phenotypes considered. The solid, larger bars depict the total number of phenotypes considered for that domain, whereas the more transparent, shorter bars depict the total number of phenotypes retained for analyses.

Our multivariate GWAS of 11 psychiatric diagnoses/phenotypes replicated Grotzinger et al.^23^ and characterized the genomic architecture of four correlated psychopathology factors (**Fig. 2**; **Extended Data Figs. 1-3; Supplemental Tables S1-S5**; **Methods**): **1) *Compulsive*** (n_effective_=39,364; i.e., OCD, Anorexia, and Tourette Syndrome; 11 genome-wide significant [GWS] loci), **2) *Psychotic*** (n_effective_=160,932; i.e., Schizophrenia, Bipolar Disorder, and Problematic Alcohol Use; 224 GWS loci), **3) *Neurodevelopmental*** (n_effective_=37,665; i.e., ADHD, Autism Spectrum Disorder, PTSD, MDD, Tourette Syndrome, and Problematic Alcohol Use; 14 GWS loci), and **4) *Internalizing* (**n_effective_=432,638; i.e., MDD, GAD, PTSD, and Problematic Alcohol Use; 7 GWS loci).

**Figure 2.**
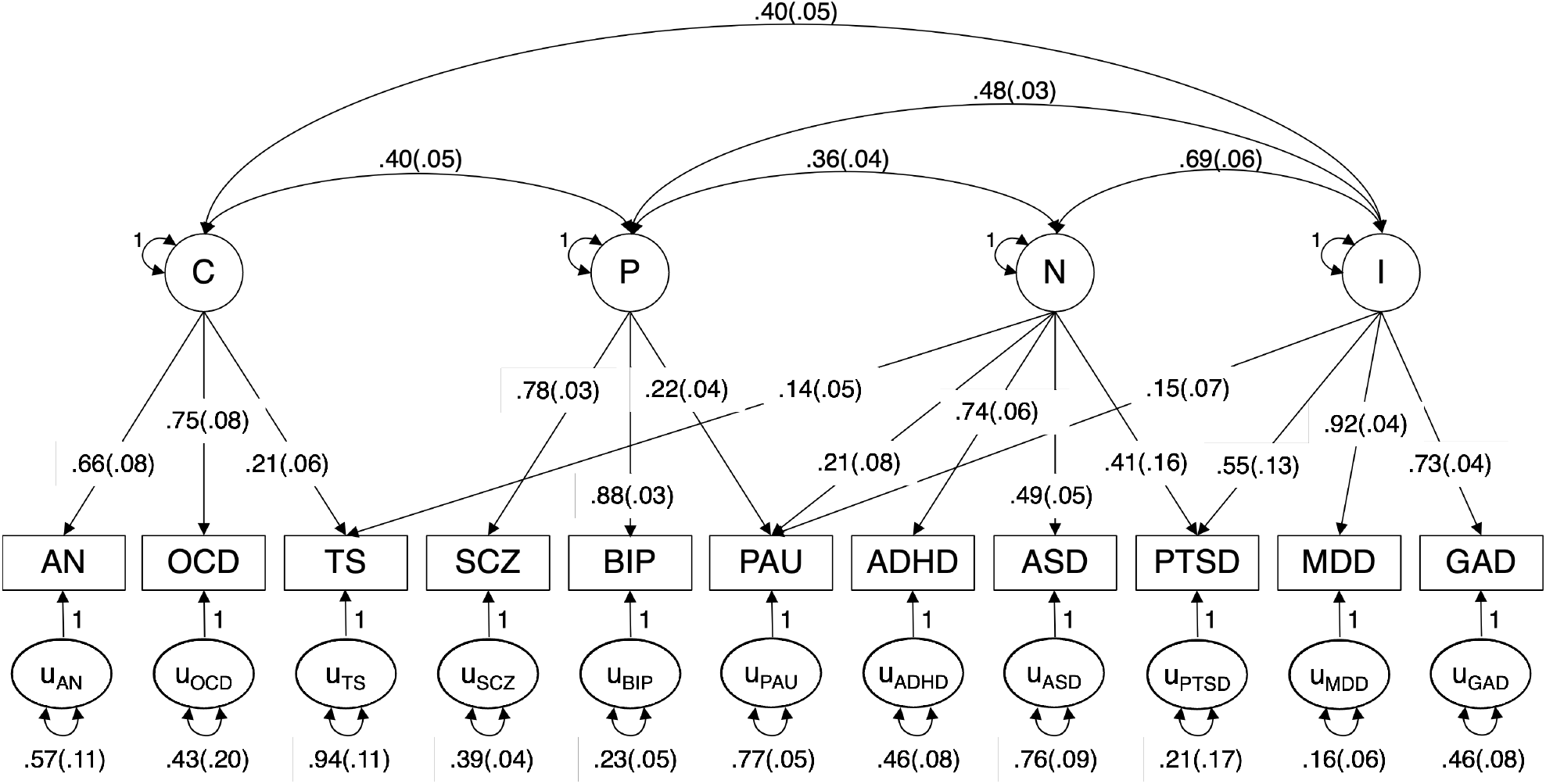
Confirmatory Factor Analysis of 11 Psychiatric Phenotypes. Confirmatory factor analysis model. Loadings and covariances are standardized. Psychiatric phenotype manifest indicators are depicted in rectangles; inferred common genetic components are represented by circles. AN = Anorexia Nervosa; OCD = Obsessive-Compulsive Disorder; TS = Tourette Syndrome; SCZ = Schizophrenia; BIP = Bipolar Disorder; PAU = Problematic Alcohol Use; ADHD = Attention-Deficit/Hyperactivity Disorder; ASD = Autism Spectrum Disorder; PTSD = Post-Traumatic Stress Disorder; MDD = Major Depressive Disorder; GAD = Generalized Anxiety Disorder.

We conducted a PheWAS of polygenic risk scores (PRS) for these factors (i.e., Compulsive, Psychotic, Neurodevelopmental, and Internalizing PRS) with baseline (n_analytic_ up to 5,556 ; 9.93±0.63 years; 47.0% female) and 2-year follow-up (FU2; n_analytic_ up to 5,048; 12.02±0.66 years; 46.93% girls) ABCD data (**Table 1**). Bonferroni and False Discovery Rate (FDR) correction were applied to all associations, separately for each PRS (i.e., 1,269 and 1,694 at baseline and FU2, respectively). Analyses revealed multiple associations with Neurodevelopmental PRS (n_fdr_=637-702; n_Bonferroni_=187-211; **Fig. 3a**) and Internalizing PRS (n_fdr_=601-687; n_Bonferroni_=122-173; **Fig. 3b**) with few associations identified for Psychotic PRS (n_fdr_=7; n_Bonferroni_=1; **Fig. 3d**) and none identified for Compulsive PRS **(Fig. 3d; Table 2**).

**Table 1.**
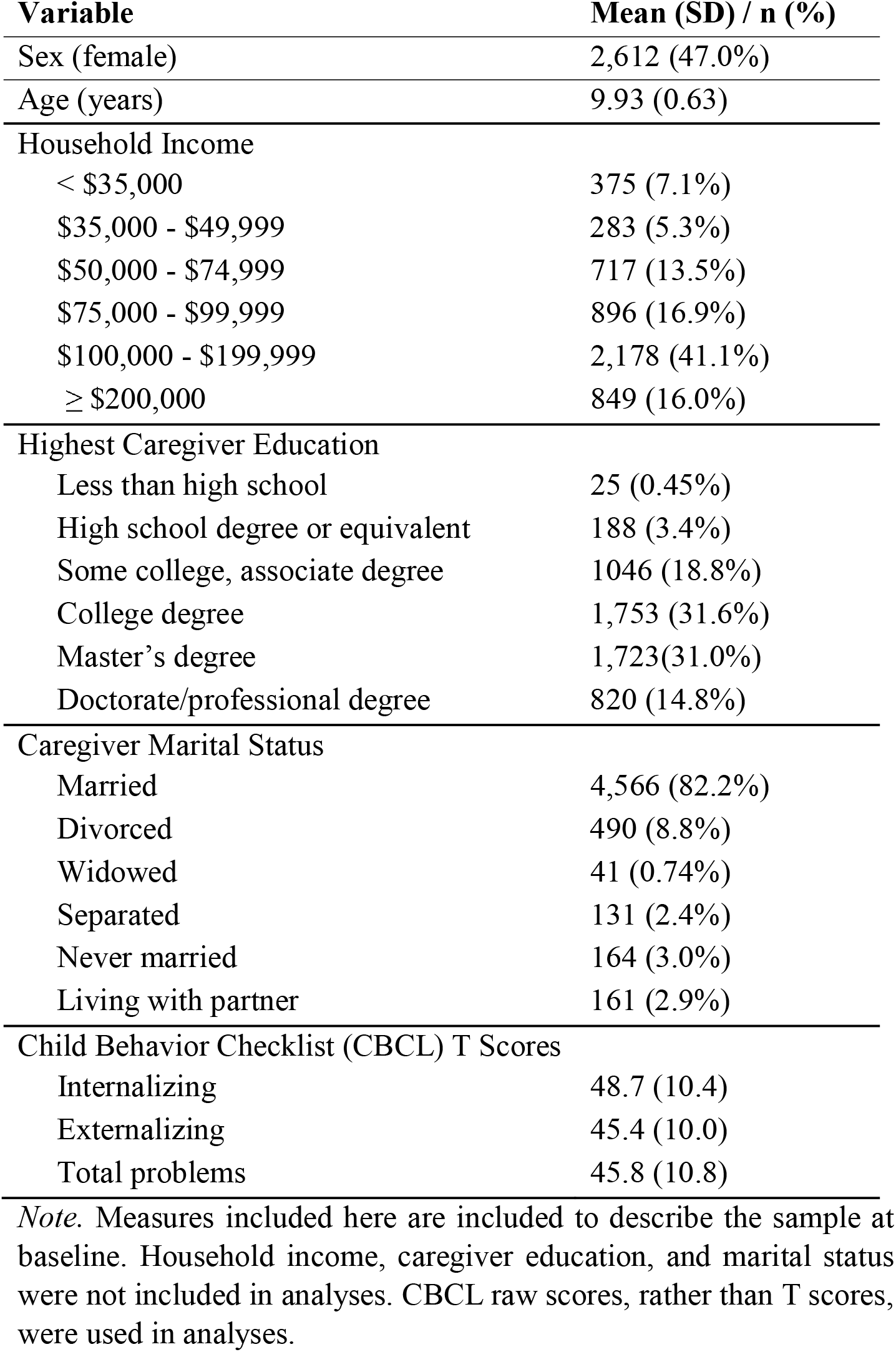
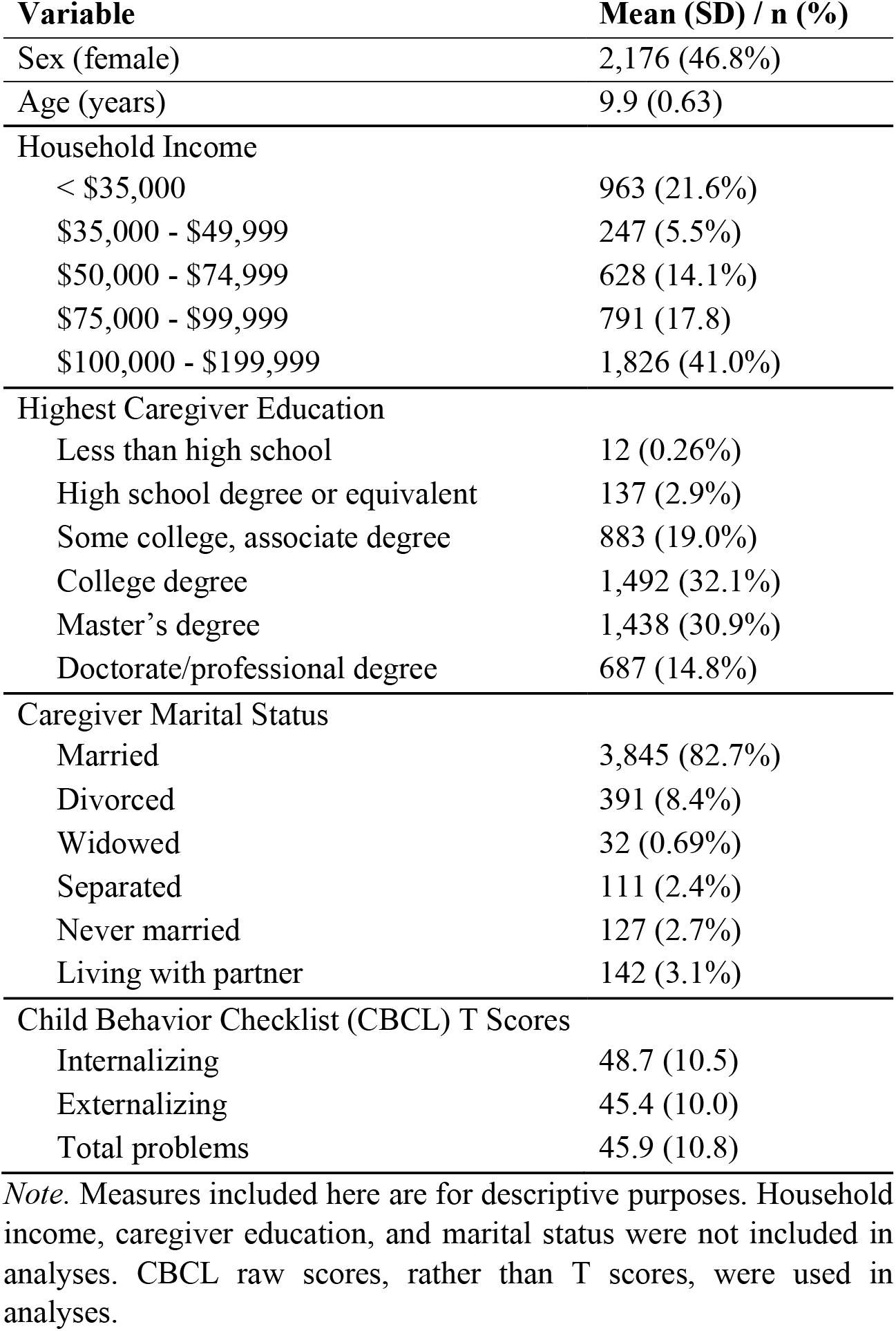
Sample characteristics.

**Figure 3.**
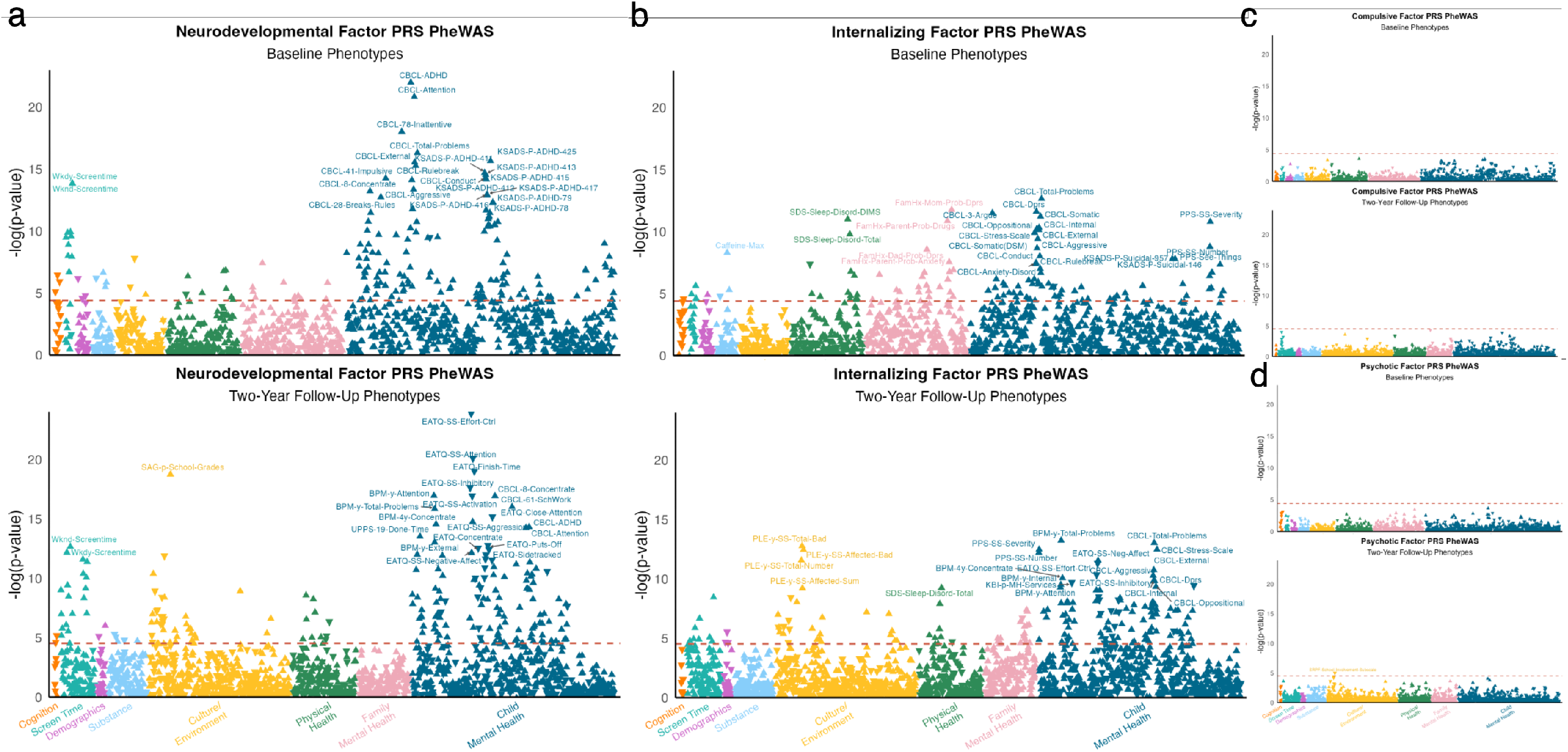
Manhattan plots of PheWAS results…. **a**. Neurodevelopmental PRS-PheWAS results (top: Baseline; bottom: FU2). **b**. Internalizing PRS-PheWAS results (top: Baseline; bottom: FU2). **c**. Compulsive PRS-PheWAS results (top: Baseline; bottom: FU2). **d**. Psychotic PRS-PheWAS results (top: Baseline; bottom: FU2). The x-axes represent phenotypes, and the y-axis represent the -log_10_ of the uncorrected *p* values of associations between each PRS and each phenotype. Each dot represents one phenotype, with the colors indicating categories and triangle direction indicating directionality of effect. The dashed lines indicate the threshold to survive Bonferroni correction. Full results can be found in **Supplemental Tables S11-S12**.

**Table 2.** Summary of significant associations.

|  |  | **Baseline** |  |  |  |  |  |  |  |
| --- | --- | --- | --- | --- | --- | --- | --- | --- | --- |
|  | Total | Compulsive |  | Psychotic |  | Neurodevel. |  | Internalizing |  |
|  |  | FDR | Bonf | FDR | Bonf | FDR | Bonf | FDR | Bonf |
| Child MH | 640 | 0 (0%) | 0 (0%) | 0 (0%) | 0 (0%) | 403 (63%) | 144 (23%) | 354 (55%) | 74 (12%) |
| Fam MH | 239 | 0 (0%) | 0 (0%) | 0 (0%) | 0 (0%) | 97 (41%) | 11 (4.6%) | 128 (54%) | 26 (11%) |
| Physical Health | 172 | 0 (0%) | 0 (0%) | 0 (0%) | 0 (0%) | 53 (31%) | 9 (5.2%) | 59 (34%) | 13 (7.6%) |
| Environment | 113 | 0 (0%) | 0 (0%) | 0 (0%) | 0 (0%) | 36 (32%) | 3 (2.7%) | 17 (15%) | 0 (0%) |
| Demographics | 27 | 0 (0%) | 0 (0%) | 0 (0%) | 0 (0%) | 10 (37%) | 3 (11%) | 10 (37%) | 1 (3.7%) |
| Substance | 46 | 0 (0%) | 0 (0%) | 0 (0%) | 0 (0%) | 15 (33%) | 4 (8.7%) | 11 (24%) | 3 (6.5%) |
| Screen Time | 18 | 0 (0%) | 0 (0%) | 0 (0%) | 0 (0%) | 13 (72%) | 10 (56%) | 14 (78%) | 4 (22%) |
| Cognition | 14 | 0 (0%) | 0 (0%) | 0 (0%) | 0 (0%) | 10 (71%) | 3 (21%) | 9 (64%) | 1 (7.1%) |
|  |  | **Two-Year Follow-Up** |  |  |  |  |  |  |  |
|  | Total | Compulsive |  | Psychotic |  | Neurodevel. |  | Internalizing |  |
|  |  | FDR | Bonf | FDR | Bonf | FDR | Bonf | FDR | Bonf |
| Child MH | 636 | 0 (0%) | 0 (0%) | 2 (0.3%) | 0 (0%) | 300 (47%) | 134 (21%) | 315 (50%) | 108 (17%) |
| Fam MH | 158 | 0 (0%) | 0 (0%) | 1 (0.6%) | 0 (0%) | 45 (28%) | 0 (0%) | 98 (62%) | 19 (12%) |
| Physical Health | 199 | 0 (0%) | 0 (0%) | 0 (0%) | 0 (0%) | 67 (34%) | 11 (5.5%) | 66 (33%) | 9 (4.5%) |
| Environment | 441 | 0 (0%) | 0 (0%) | 3 (0.7%) | 1 (0.2%) | 162 (37%) | 39 (8.8%) | 125 (28%) | 26 (5.9%) |
| Demographics | 26 | 0 (0%) | 0 (0%) | 0 (0%) | 0 (0%) | 11 (42%) | 2 (7.7%) | 7 (27%) | 2 (7.7%) |
| Substance | 119 | 0 (0%) | 0 (0%) | 0 (0%) | 0 (0%) | 53 (45%) | 5 (4.2%) | 20 (17%) | 0 (0%) |
| Screen Time | 107 | 0 (0%) | 0 (0%) | 1 (0.9%) | 0 (0%) | 60 (56%) | 19 (18%) | 54 (50%) | 9 (8.4%) |
| Cognition | 8 | 0 (0%) | 0 (0%) | 0 (0%) | 0 (0%) | 5 (63%) | 1 (13%) | 2 (25%) | 0 (0%) |

*Note.* Summary of the number of significant associations per domain, at each wave of assessment. The total number of included phenotypes per domain are represented in the second column. The number and percent (of the total number of phenotypes in that domain) of significant associations (FDR- and Bonferroni-corrected p-values) are depicted. Bonf. = Bonferroni. Neurodevel. = Neurodevelopmental. Percentages are rounded to two digits.

### Neurodevelopmental PRS-PheWAS

Neurodevelopmental PRS was significantly associated with ≥187 phenotypes at each timepoint (n_baseline-fdr_=637; n_baseline-Bonferroni_=187; n_FU2-fdr_=702; n_FU2-Bonferroni_=211; **Table 2; Fig. 3a; Extended Data Figs. 4-5, Supplemental Tables S11-S12**). The child mental health and screen time domains were the most frequently associated with Neurodevelopmental PRS across both waves (**Table 2**), with proximal (i.e., those mirroring discovery GWAS phenotypes) neurodevelopmental phenotypes including inattention and impulsivity as well as more distal phenotypes such as total screen time, sleep problems, worse relationships with teachers, psychotic-like experiences, neighborhood crime, lower parental monitoring, and receipt of mental health services among the most significantly associated phenotypes. A sizable proportion (13-21%) of cognitive phenotypes were negatively associated with Neurodevelopmental PRS, but notably were not among the top hits and constituted a small overall number of phenotypes.

Of the 799 phenotypes assessed at both waves, 159 (n_FDR_=442) and 115 (n_FDR_=366) showed Bonferroni-corrected significant associations at baseline and FU2, respectively, of which 75 (n_FDR_=284) were consistently associated across waves. Among these phenotypes associated across waves were those reflecting child mental health (e.g., CBCL and teacher-reported Brief Problem Monitor items and scales reflecting inattentive and externalizing behaviors), special services in school, school performance, psychotic-like experiences summary scores, screen time metrics, sleep issues, and impulsive personality traits (e.g., lack of perseverance, lack of planning). Wave-specific associations (baseline: n_FDR_=349, n_Bonferroni_=112; **Supplemental Table S11**; FU2: n_FDR_=418, n_Bonferroni_=136; **Supplemental Table S12**; **Table 2**) were predominantly attributable to phenotypes only assessed at either session (e.g., prenatal exposures and family history at baseline; Autism Spectrum symptoms and youth-reported externalizing behaviors at FU2), as well as those associated with broad spectrum risk (e.g., increased youth and caregiver internalizing and broad behavior problems at baseline). As children began to progress into adolescence (FU2), Neurodevelopmental PRS showed more associations with dysfunction that typically become more evident at this age (e.g., worse school performance, conflict at school and with caregiver). Several of these associations (e.g., negative associations with liking school because they do well, feeling as smart as their peers, and school involvement) had non-overlapping 95% confidence intervals across waves (**Supplemental Table S13**), though it should be noted that some of these variables (e.g., conflict with caregiver about grades) are more frequently endorsed at FU2.

### Internalizing PRS-PheWAS

Polygenic risk for the Internalizing factor was significantly associated with ≥122 phenotypes at each timepoint (n_baseline-fdr_=601; n_baseline-Bonferroni_=122; n_FU2-fdr_=687; n_FU2-Bonferroni_=173; **Table 2; Fig. 3b; Extended Data Figs. 4-5, Supplemental Tables S11-S12**). The child mental health, family mental health, and screen time domains were most frequently associated with Internalizing PRS, with Bonferroni-corrected significant associations ranging from 8.4% (screen time at FU2) to 22% (screen time at baseline) of assessed phenotypes. The other domains showed less frequent associations, with between 0% (culture/environment at baseline, and substance-related and cognition phenotypes at FU2) and 7.7% (Demographics at FU2) of associations surviving Bonferroni correction (**Table 2**). Top associations included proximal internalizing phenotypes, such as depressive and stress-related symptoms, as well as more distal phenotypes including total behavior problems, psychotic-like experiences, sleep difficulties, and externalizing behaviors.

Of the 799 phenotypes assessed at both waves, 98 (n_FDR_=414) and 100 (n_FDR_=386) were significantly (Bonferroni) associated with Internalizing PRS at baseline and FU2, respectively, of which 46 (n_FDR_=275) were consistently associated across both waves. Featured among these phenotypes were those reflecting internalizing, externalizing, and total behavior problems; psychotic-like experiences, sleep disturbances, and school difficulty and special services. Associations specific to each wave (baseline: n_FDR_=326, n_Bonferroni_=76; **Supplemental Table S11**; FU2: n_FDR_=412, n_Bonferroni_=172; **Supplemental Table S12**; **Table 2**) largely reflected phenotypes assessed only at one timepoint (e.g., family history and prenatal exposures at baseline; youth-reported behavior problems, items on the Early Adolescent Temperament Questionnaire, stressful life events at FU2), as well as additional school-related risk factors (e.g., drop in grades, detention/suspension, self-reported dislike of school and feeling less smart than peers) and caregiver-reported mood swings at FU2. As with Neurodevelopmental PRS, 95% confidence intervals for several of these associations were non-overlapping across waves, with stronger associations observed at FU2 including both feeling less smart than peers and psychotic-like experience items, which may overlap in content with social anxiety characteristics (i.e., suddenly not trusting others who seem to be watching or talking about them, difficulty figuring out how to say something so others can understand; **Supplemental Table S13**).

### Compulsive PRS-PheWAS

Polygenic risk for the Compulsive factor was not significantly associated with any phenotypes at either wave following FDR and Bonferroni correction (**Fig. 3c**; **Extended Data Figs. 4-5, Supplemental Tables S11-S12**). Nominally significant associations (i.e., *p*<0.05) included past OCD diagnosis and symptoms at baseline as well as negative associations with caregiver self-reported poor diet, time spent watching TV on the weekend, and measured weight and waist circumference at FU2.

### Psychotic PRS-PheWAS

Polygenic risk for the Psychotic factor was significantly associated with lower school involvement at FU2 (n_baseline-fdr_=0; n_baseline-Bonferroni_=0; n_FU2-fdr_=7; n_FU2-Bonferroni_=1; **Fig. 3d; Extended Data Figs. 4-5, Supplemental Tables S11-S12**). Nominally significant associations included functional impairment due to compulsions, caregiver history with mania, and negative associations with child-reported school factors (e.g., feeling as smart as peers, liking school because they are doing well**). Figure 4** summarizes primary PheWAS results.

**Figure 4.**
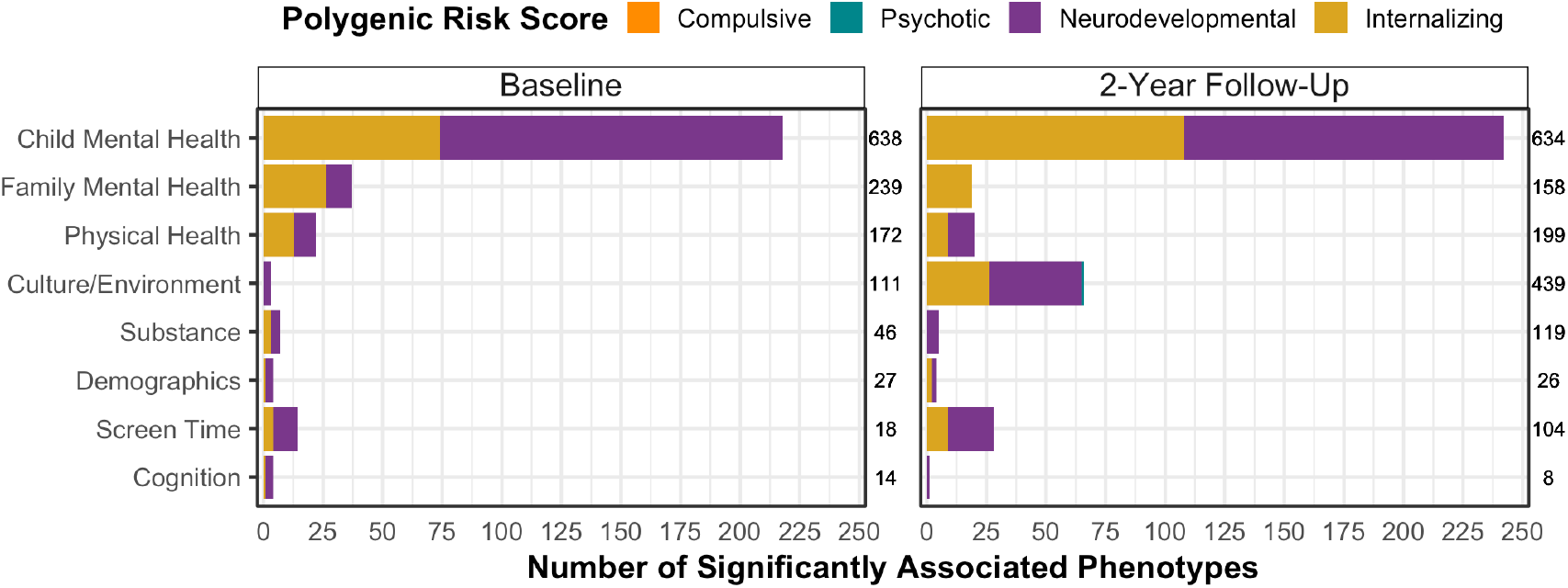
Summary of PheWAS findings. Summary of phenotypes significantly associated with each PRS, grouped by phenotype domain. The left y-axis reflects the phenotype domains. The right y-axes show the total number of phenotypes analyzed per domain. The x-axis denotes the number of phenotypes significantly associated with each PRS.

### Imaging Analyses

Compulsive PRS was significantly and positively associated with three imaging metrics: left precentral volume and surface area, and right transverse temporal gyrus (βs ≥0.032, *p*s≤5.6e-04). Psychotic PRS showed significant negative associations with right pars orbitalis thickness, right ventral diencephalon volume, and fractional anisotropy of the corpus callosum (βs≤-0.017, *p*s≤1.1e-03). Neurodevelopmental PRS was broadly negatively associated with global volume metrics (i.e., total, total right, and total left hemisphere cortical volume, whole brain volume, supratentorial volume), right mean thickness, and 4^th^ ventricle volume (βs≤-0.037, *p*s≤2.6e-03). Internalizing PRS was not associated with any imaging metrics following Bonferroni correction (**Supplemental Tables S14-S21; Extended Data Figs. 6-10**).

### Indirect Effects Analyses

To explore whether brain metrics may indirectly link PRS to two-year follow-up phenotypes, associations between each of the significantly associated imaging measures and each of the significantly associated phenotypes at follow-up were examined (i.e., Psychotic PRS: 3 imaging metrics * 1 phenotype; Neurodevelopmental PRS: 5 imaging metrics * 211 phenotypes). None of the imaging metrics associated with Psychotic PRS were significantly linked to school involvement, the single phenotype whose association with Psychotic PRS survived Bonferroni correction. For Neurodevelopmental PRS, 47-51 FU2 phenotypes showed significant associations with total cortical volume, supratentorial volume, and whole brain volume (|βs|≤0.07, *p*s≤4.5e-04) following Bonferroni correction within each imaging metric. The strongest associations were between all three volume indices and indicators of school performance, with smaller volumes being linked to worse performance. Other phenotypes negatively associated with brain volumes included CBCL externalizing symptoms, issues with concentration, symptoms of mania, total weekday screen time, and being on Medicaid insurance. Path analysis models including the direct effect of PRS on follow-up phenotype in addition to the indirect effect of PRS on follow-up phenotype via cortical volume were fit. Only one association survived Bonferroni correction (for 47-51 tests, depending on metric): Total cortical volume indirectly linked Neurodevelopmental PRS to grades in school. Neurodevelopmental PRS was negatively associated with total cortical volume (β=-0.056, 95% CI [-0.087, -0.026]), which was negatively associated with worse grades in school (β=-0.14, 95% CI [-0.18, -0.11]), and indirectly linked Neurodevelopmental PRS to worse grades in school (direct effect β=0.012; indirect effect β=0.0081, *p*=8.9e-04; **Figure 5**). Although the majority of other indirect effects were significant at the 0.05 level, none survived correction for the total number of tests. All results are represented in **Supplemental Tables S22-S23**.

**Figure 5.**
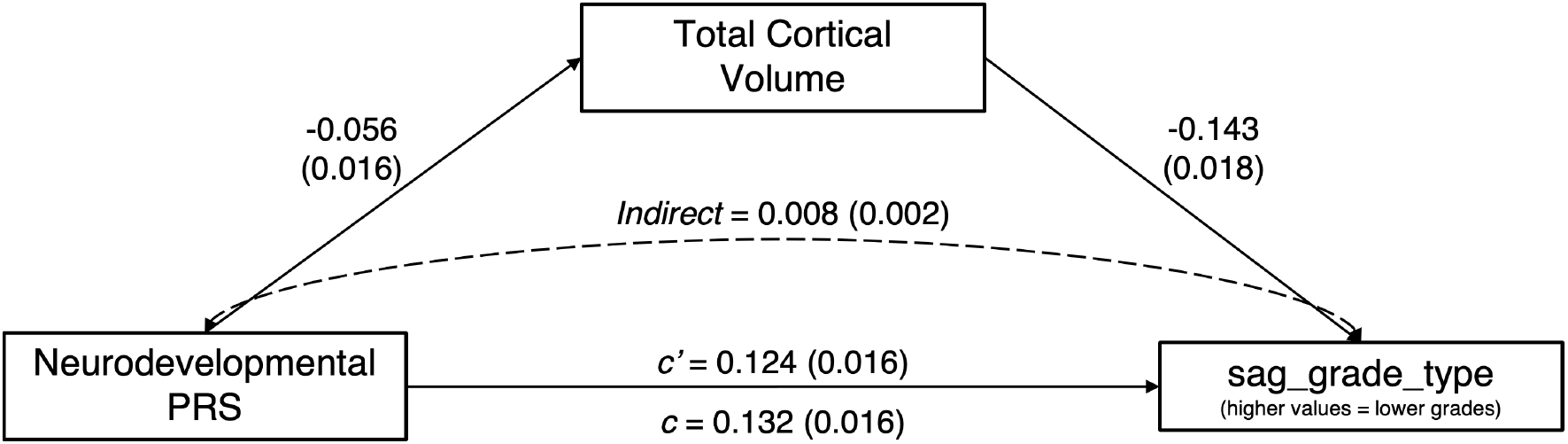
Indirect effect of Neurodevelopmental PRS on school grades through cortical volume. All estimates are standardized and statistically significant (*p*_*BONFERONNI*_ < 0.05). *c* = total effect; *c’* = direct effect.

## DISCUSSION

Among 5,048-5,556 youth of European ancestry aged 9-13, PRS for broad spectrum Neurodevelopmental and Internalizing psychopathology were associated with numerous proximal phenotypes (e.g., inattention and impulsivity; depressive symptoms and anxiety) as well as a host of more distal variables (e.g., metrics of screen time; stressful life events). Compulsive and Psychotic PRS largely failed to predict even proximal phenotypes, with Psychotic PRS showing only one significant association with lower school involvement. Several significant associations between PRS and neuroimaging metrics emerged, such as between Neurodevelopmental PRS and global volume measures. Lower total cortical volume measured at baseline partially indirectly linked Neurodevelopmental PRS to lower grades in school assessed at two-year follow-up. This study builds upon prior PRS-PheWAS by leveraging PRS of higher order latent factors representing phenotypes across the psychiatric diagnostic spectrum, an approach that enhances the power of PRS to explain meaningful variation in proximal phenotypes and better captures the highly polygenic nature of psychopathology risk compared to individual disorder PRS.

A first key finding from this work is that both Neurodevelopmental and Internalizing PRS were associated with a range of neurodevelopmental and internalizing phenotypes across waves, far more so than Compulsive and Psychotic PRS. In particular, across both waves, Neurodevelopmental PRS was consistently and strongly associated with behaviors and symptoms of inattention, impulsivity, and externalizing disorders as reported by caregivers, teachers, and the youth themselves. For Internalizing PRS, top hits included depressive, internalizing, and stress-related symptoms, as well as family history of depression and anxiety. These results provide validation to the multivariate GWAS PRS-PheWAS approach to detecting and replicating associations with proximal phenotypes.

Neurodevelopmental and Internalizing PRS exhibited both dissociable associations and substantial convergence, with 33%-57% of significant hits at both waves shared across these PRS. Top hits with Internalizing PRS were broader in nature, including associations with general and externalizing psychopathology symptoms (e.g., on the CBCL and Brief Problem Monitor measures), psychotic-like experiences, sleep disorder issues, and stressful life events of similar magnitude to associations with internalizing symptoms. Although Neurodevelopmental PRS was linked to multiple phenotypes across these domains, top hits were more constrained to Neurodevelopmental disorder symptoms and behaviors related to impulsivity and impaired attention. This pattern of results converges with phenotypic findings in the ABCD sample that stressful life events are more strongly related to internalizing symptoms, whereas contextual variables related to externalizing behaviors, such as school factors, are more strongly related to externalizing symptoms^13^. At the same time, the substantial overlap in associations aligns with previous findings in this sample showing that latent neurodevelopmental and internalizing factors are moderately correlated with one another, and, further, that general and domain-specific factors together explain the most variance in psychopathology symptoms, school performance, and measures of cognitive ability^14^. Overall, the broad convergence across genetic liability for psychopathology spectra not only includes psychopathology phenotypes but also phenotypes overlapping with cognition and physical health.

Beyond demonstrating that many previously identified phenotypic associations hold at the level of polygenic risk for higher order psychopathology factors in children prior to the typical expression of psychiatric disorder, the present study further highlights associations with more distal phenotypes that may pave the way for future etiologic understanding. Several substance-related phenotypes were associated with Neurodevelopmental PRS, including prenatal tobacco exposure, ease of obtaining cigarettes and marijuana, and peer approval of use of cigarettes and marijuana. These findings add to prior work showing that ADHD PRS is linked to tobacco use disorder in adults^15^ and that “deviant” peer affiliation may link ADHD to future substance use^16^. These associations may be in part due potential gene-environment correlation between parental liability to Neurodevelopmental disorders and raising children in environments with increased access to substances. The large number of associations between Neurodevelopmental PRS and metrics of screen time replicates a recent finding linking ADHD PRS to total screen time use in the ABCD sample^11^ and offers further insight into specific screen media behaviors. In the present analyses, there was evidence consistent with the idea that genetic vulnerability for neurodevelopmental concerns may increase susceptibility to the rewarding aspects of screen media (e.g., screen time as a source of motivation) as well as increase aspects of problematic use (e.g., struggling to limit screen time).

The numerous associations between Internalizing PRS and sleep difficulties, psychotic-like experiences, and stressful life events and peer victimization also highlight fruitful avenues for further exploration. Diagnostic criteria for depression and anxiety include sleep difficulties, which, according to the present analyses, may be particularly salient and modifiable risk factors for internalizing disorders. Psychotic-like experiences have previously been found to show strong associations with broad psychopathology in the ABCD study sample, including internalizing symptoms, and the present findings strengthen the notion that psychotic-like experiences may constitute a broad manifestation of psychopathology risk in childhood^17^. Furthermore, associations between internalizing symptom genetic liability and stressful life events can be interpreted as being consistent with gene-environment correlation and stress generation theories and evidence that parental depression may contribute to stress experienced by children^18,19^. Associations between Internalizing PRS and self-reported peer victimization phenotypes (e.g., peer gossip and threats to reputation) are also consistent with the theory that relational stressors may be experienced relatively more frequently by those at risk for internalizing symptoms^18^.

By contrast, PRS for Compulsive and Psychotic disorders showed zero and one significant associations, respectively. There are several non-mutually exclusive explanations for this pattern of findings. *First*, the psychiatric phenotypes comprising the Compulsive and Psychotic factors (e.g., OCD, Anorexia, Schizophrenia, Bipolar Disorder) are generally less prevalent than disorders such as ADHD, Generalized Anxiety, and Major Depression^1,20^. As such, the ABCD sample, which was recruited to be representative of the United States 9–10-year-old population, may be insufficiently enriched for Compulsive and Psychotic disorders for associations to be detected with genetic liability. *Second*, and relatedly, disorders such as Schizophrenia and Bipolar Disorder in particular, and to a lesser degree OCD and Eating Disorders, typically manifest later than ADHD and anxiety. It is possible that associations with Compulsive and Psychotic PRS may arise as the ABCD sample ages. Notably, the relative lack of findings for Psychotic PRS mirrors prior work in the ABCD sample that found that Schizophrenia PRS were not significantly associated with continuously measured psychotic-like experiences, whereas cross-disorder PRS were^21^. It is possible that phenotypic expression of underlying genetic liability to these domains may be observed as the sample ages, which would converge with a recent PheWAS documenting associations between PRS for Schizophrenia and psychotic symptoms in adults^15^.

A second set of analyses examined associations between each PRS and brain structure and resting state functional connectivity, adding to the emerging neurogenetics literature. Findings for Compulsive PRS include a positive association with right transverse temporal cortex surface area, notable in its direction given prior evidence in the ENIGMA OCD sample (and lack of association in the ENIGMA study of children and adolescent OCD), with the present sample perhaps pointing to the influence of maturational changes specific to middle childhood .^22^ OCD has been previously linked to increased intra-frontoparietal network connectivity^23,24^, as in the present study, and may reflect over control and excessive monitoring tendencies typical of compulsive disorders. Psychotic PRS was associated with lower par orbitalis thickness, lower ventral diencephalon volume, and lower fractional anisotropy of the corpus callosum, consistent with previous research on clinical high risk for psychosis (though pars orbitalis thickness may be associated especially with more chronic diseases stages)^25–27^. Neurodevelopmental PRS was negatively associated with global gray matter volume, as well as right cortical thickness, whereas Internalizing PRS showed no significant associations with neural metrics. This pattern aligns with recent work showing widespread negative associations between externalizing, but not internalizing, symptoms and cortical and subcortical regional gray matter volume, although notably the majority of significant associations did not survive covarying for total intracranial volume^28^.

Given emerging evidence that brain structure may mechanistically link genetic liability to phenotypic expression ^e.g.,9,29^, we probed whether baseline neural metrics assessed significantly associated with PRS indirectly link said PRS with significantly associated phenotypes assessed at two-year follow-up. We found that smaller total cortical volume indirectly linked Neurodevelopmental PRS to poorer grades in school, extending prior work showing that brain volume is positively associated with educational performance^30^. Global gray matter structural alterations may be one mechanism through which polygenic liability to neurodevelopmental disorders may impact school performance, and although future analyses should explore putative environmental and other mechanisms. It should also be noted that generally small genetic correlations between psychopathology and neural metrics may constrain power to detect these associations^31^.

### Limitations

The major limitation of the present study is the exclusion of genetic ancestry groups other than European from analyses. Although this constraint is attributable to the lack of available GWAS summary statistics on the 11 psychiatric phenotypes comprising the multivariate GWAS in other genetic ancestry groups and the poor predictive ability of PRS across genetic ancestry ^32^, the applicability of these findings only to those of European genetic ancestry has the potential to increase disparities in healthcare interventions and access to information based on ancestry. All findings described above should be interpreted with caution, as they may have poor generalizability beyond Americans of European genetic ancestry.

In addition, these latent higher order genetic psychopathology factors may be missing important diagnoses, such as social anxiety, conduct, and oppositional defiant disorders, as GWAS on these disorders are either not available or are severely underpowered. The Compulsive factor consisted of the three disorders with lowest Ns, which may have weakened power for PRS development and discovery. It is worth noting, however, that power is not the only driver of discovery, as three of the largest samples loaded on the psychotic factor, which arguably had more power to train PRS but was significantly associated with fewer phenotypes than the Neurodevelopmental or Internalizing PRS. Numerous associations were observed in the context of a relatively small Neurodevelopment GWAS effective sample size (NEffective=37,665), perhaps reflecting the developmentally normative nature of neurodevelopmental phenotypes observed in this age group. Further, although we elected to use strict p-value correction (i.e., Bonferroni) given the large number of tests and exploratory nature of analyses, this method may be overly stringent in ignoring the large pattern of correlation among phenotypes, and therefore we also provide FDR correction estimates in all results tables. Finally, although the present analyses do not extend into peak ages of psychiatric risk for most disorders, it is important to identify phenotypes in childhood that may represent critical signatures of genetic risk for psychopathology.

## Conclusions

This study builds on prior psychiatric PRS-PheWAS^15,33,34^ by interrogating associations between PRS derived from higher order latent factors representing phenotypes across the diagnostic spectrum in a sample of youth. The young age of the sample and related low levels of diagnosable psychiatric disorders enabled the identification of putative risk factors associated with genetic liability largely prior to disorder onset. Further, in contrast to the Electronic Health Records approach of many PheWAS studies, the present study leveraged deeply characterized phenotypes encompassing both threshold and subthreshold psychopathology symptoms as well as a host of other domains not typically included in EHR data (e.g., brain structure, screen time, environmental exposures, multiple reporters and modes of assessment). The present study provides support for the power of agnostic methods to improve understanding of the etiopathogenesis of psychopathology in youth.

## Data Availability

All data produced in the present study are available upon reasonable request to the authors.
Genome-wide association study summary statistics will be posted in a link here following medRxiv approval.

## Acknowledgements

SEP, NRK, DMB, and RB developed the research questions. SEP, AJG, NRK, IN, LB, ISH, and NME cleaned the phenotypic data. SEP, SC, AJG, and NRK conducted analyses. SEP and NRK drafted the manuscript. SC, AJG, ISH, ECJ, ASH, DAAB, NME, DB, and RB provided critical revision of the manuscript for important intellectual content. SEP and NRK had full access to all the data in the study and take responsibility for the integrity of the data and the accuracy of the data analysis.

## Code availability

Code for Genomic SEM and PheWAS analyses are located on GitHub: **https://github.com/sepaul1124/ABCD-PHEWAS**

---

**Extended Data Figure 1.**
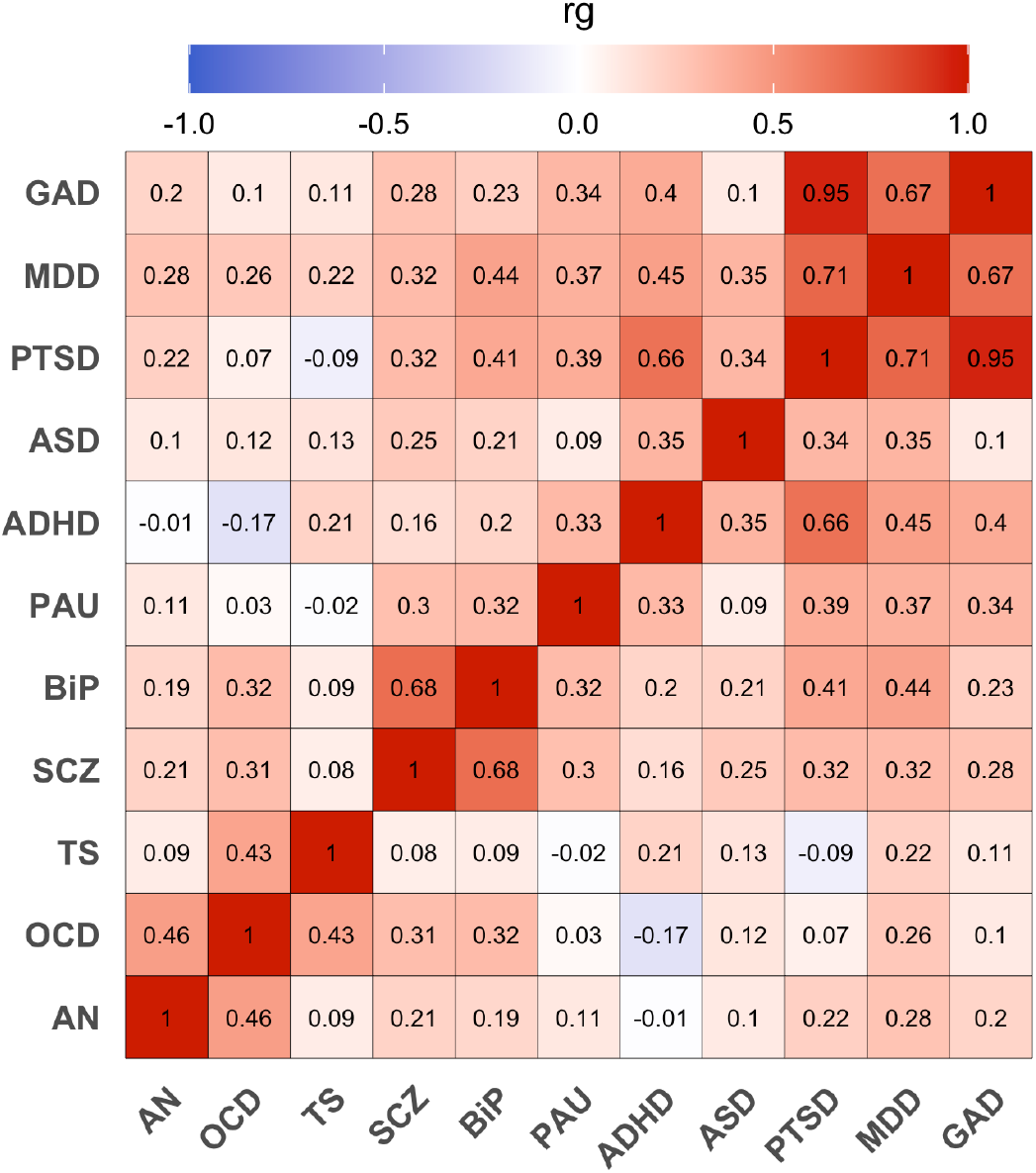
Genetic Correlations Among 11 Psychiatric Phenotypes. AN = Anorexia Nervosa; OCD = Obsessive-Compulsive Disorder; TS = Tourette Syndrome; SCZ = Schizophrenia; BIP = Bipolar Disorder; PAU = Problematic Alcohol Use; ADHD = Attention-Deficit/Hyperactivity Disorder; ASD = Autism Spectrum Disorder; PTSD = Post-Traumatic Stress Disorder; MDD = Major Depressive Disorder; GAD = Generalized Anxiety Disorder. Genetic correlations were estimated using the ldsc() function in the GenomicSEM R package.

**Extended Data Figure 2.**
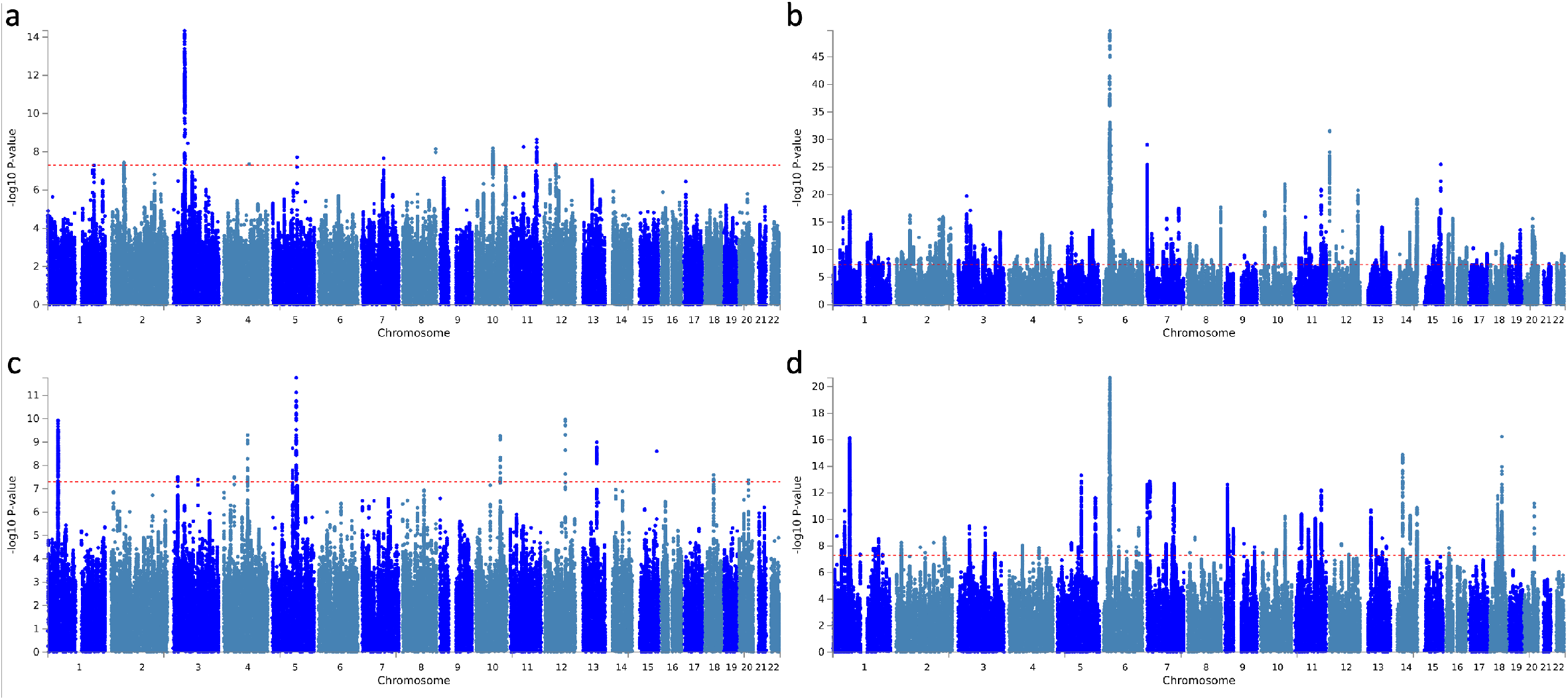
Multivariate GWAS Manhattan Plots. Multivariate GWAS Manhattan Plots for **a)** Compulsive, **b)** Psychotic, **c)** Neurodevelopmental, and **d)** Internalizing factors. Y-axes represent the negative log(10) of the SNP effect p-values. The red dotted line reflects the genome-wide significance threshold for one million tests. The x-axes show individual SNPs across chromosomes. Each dot represents a single SNP effect. Overlapping data points are not drawn. Note the different y-axes.

**Extended Data Figure 3.**
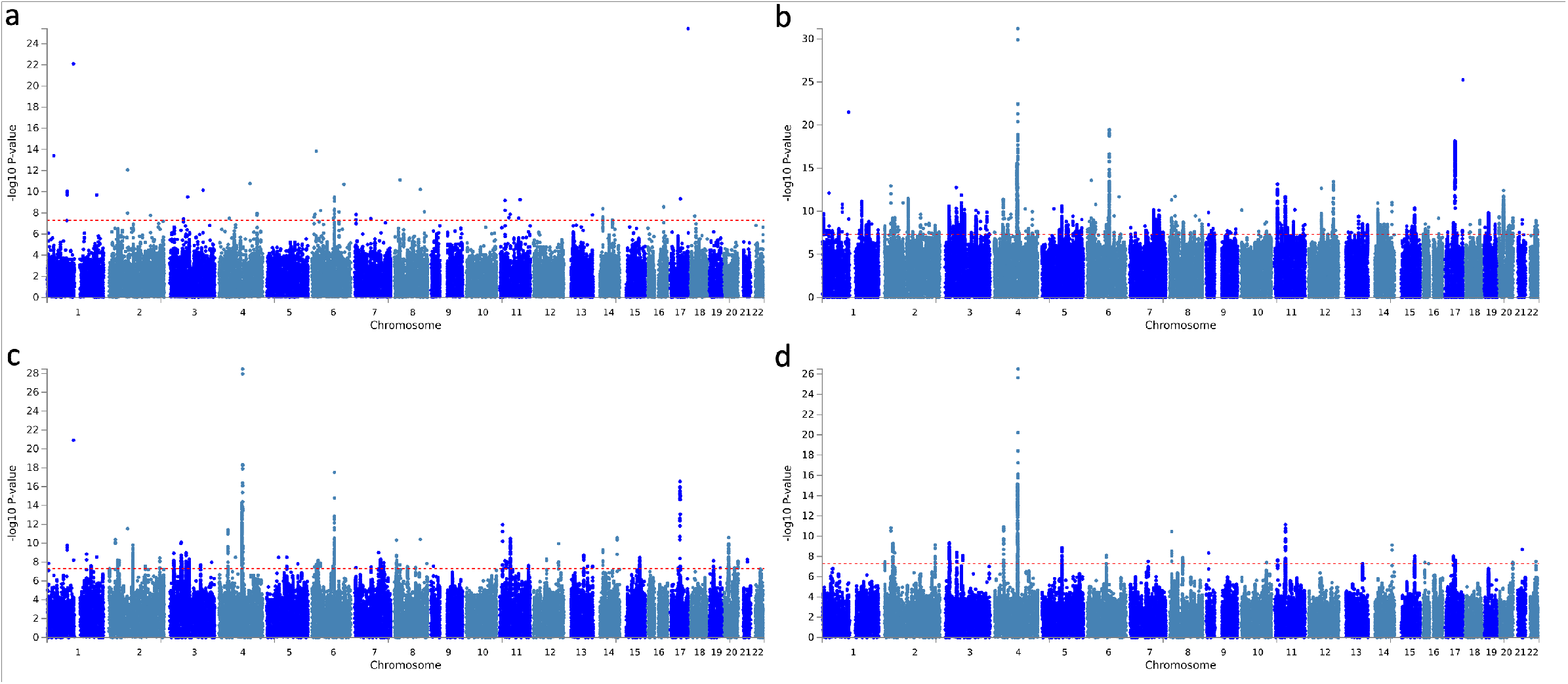
Multivariate GWAS Factor-Specific Q-SNPs Manhattan Plots. Factor-specific Q-SNP effects for **a)** Compulsive, **b)** Psychotic, **c)** Neurodevelopmental, and **d)** Internalizing factors. Y-axes represent the negative log(10) of the Q-SNP effect p-values. The red dotted line reflects the genome-wide significance threshold for one million tests. The x-axes show individual SNPs across chromosomes. Each dot represents a single Q-SNP effect. Overlapping data points are not drawn. Note the different y-axes.

**Extended Data Figure 4.**
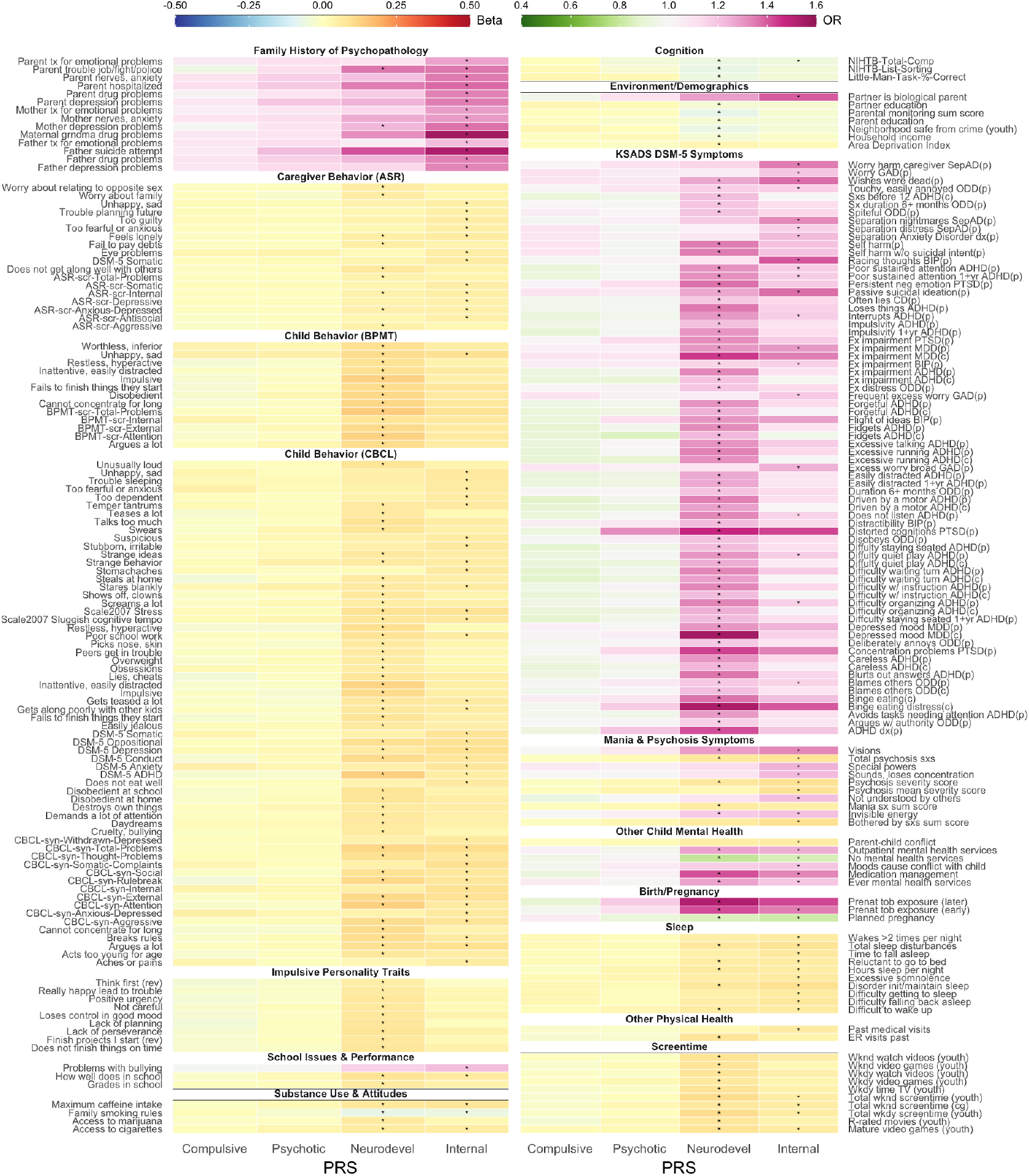
Heatmap of PheWAS baseline results. PRS are depicted along the x-axis and all phenotypes whose association with any PRS survives Bonferroni correction are labeled on the y-axes. For all phenotypes, darker colors represent larger effects. For continuous phenotypes, red colors indicate positive associations, whereas blue colors indicate negative associations. For dichotomous phenotypes analyzed with binomial generalized mixed effects models, effects sizes are represented by odds ratios (ORs), with ORs > 1 appearing pink/purple, and ORs <1 appearing green. Asterisks show Bonferroni-corrected significant associations. Phenotypes have been grouped into domains for easier interpretation.

**Extended Data Figure 5.**
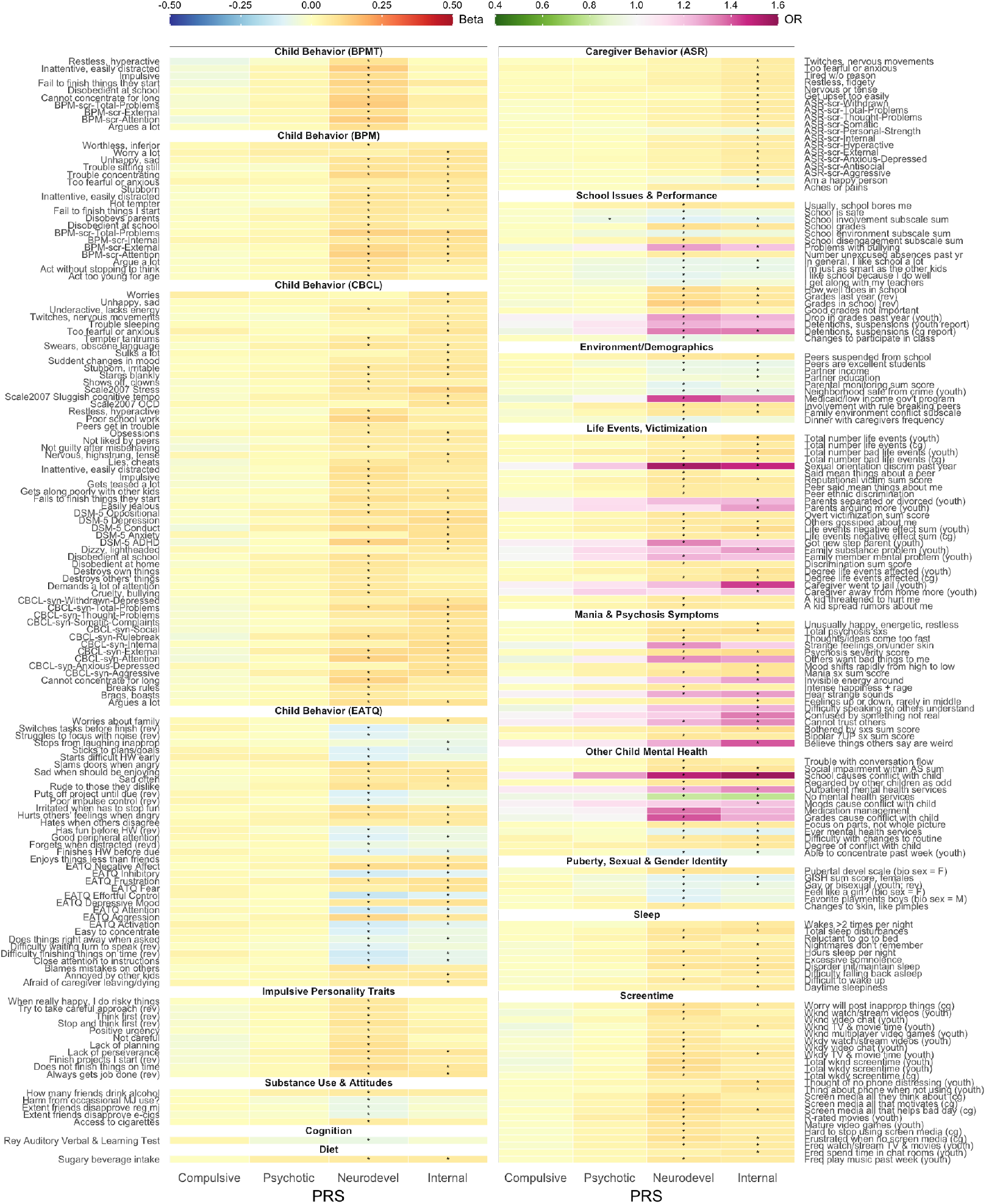
Heatmap of PheWAS two-year follow-up results. PRS are depicted along the x-axis and all phenotypes whose association with any PRS survives Bonferroni correction are labeled on the y-axes. For all phenotypes, darker colors represent larger effects. For continuous phenotypes, red colors indicate positive associations, whereas blue colors indicate negative associations. For dichotomous phenotypes analyzed with binomial generalized mixed effects models, effects sizes are represented by odds ratios (ORs), with ORs > 1 appearing pink/purple, and ORs <1 appearing green. Asterisks show Bonferroni-corrected significant associations. Phenotypes have been grouped into domains for easier interpretation.

**Extended Data Figure 6.**
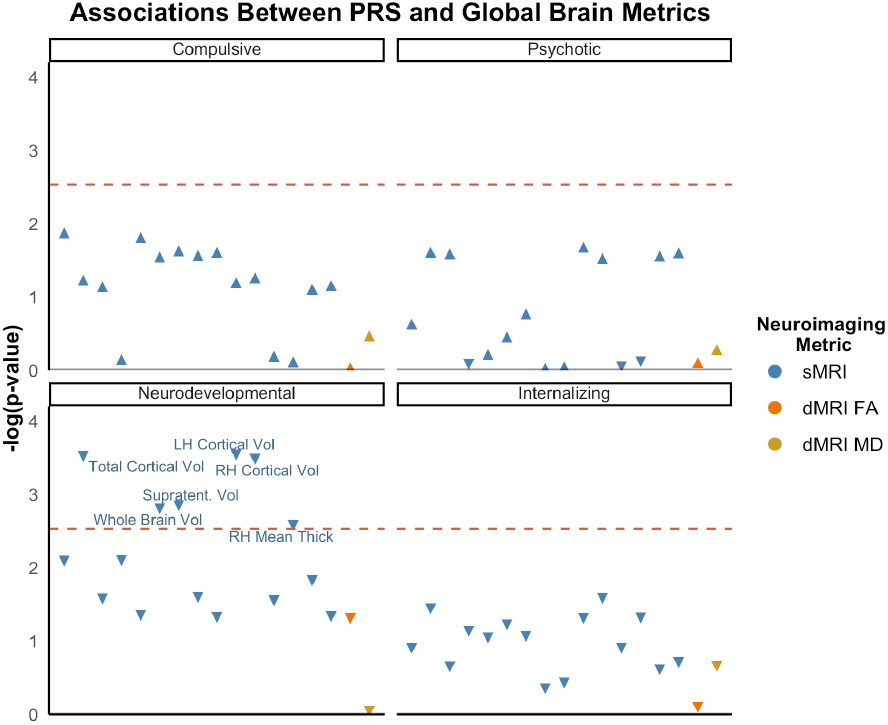
Associations between PRS and Global Brain Metrics. Associations between each PRS and global brain metrics. The x-axes represent phenotypes, and the y-axis represent the -log_10_ of the uncorrected *p* values of associations between each PRS and each phenotype. Each dot represents one phenotype, with the colors indicating categories and triangle direction indicating directionality of effect. The dashed lines indicate the threshold to survive Bonferroni correction. LH = left hemisphere; RH = right hemisphere; supratent = supratentorial; vol = volume. Full results can be found in **Supplemental Table S13**.

**Extended Data Figure 7.**
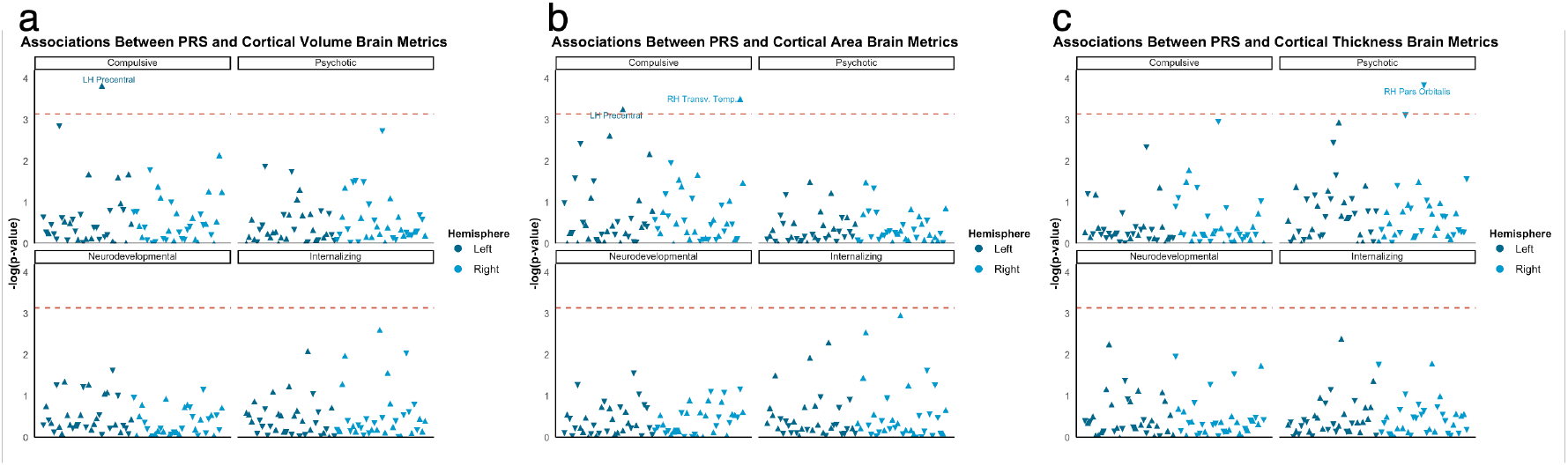
Associations between PRS and Cortical Structure. Associations between each PRS and cortical gray matter structure. **a)** Cortical Volume. **b)** Cortical Surface Area. **c)** Cortical Thickness. The x-axes represent phenotypes, and the y-axis represent the -log_10_ of the uncorrected *p* values of associations between each PRS and each phenotype. Each dot represents one phenotype, with the colors indicating hemispheres and triangle direction indicating directionality of effect. The dashed lines indicate the threshold to survive Bonferroni correction. LH = left hemisphere; RH = right hemisphere; transv = transverse; temp = temporal. Full results can be found in **Supplemental Tables S14-S16**.

**Extended Data Figure 8.**
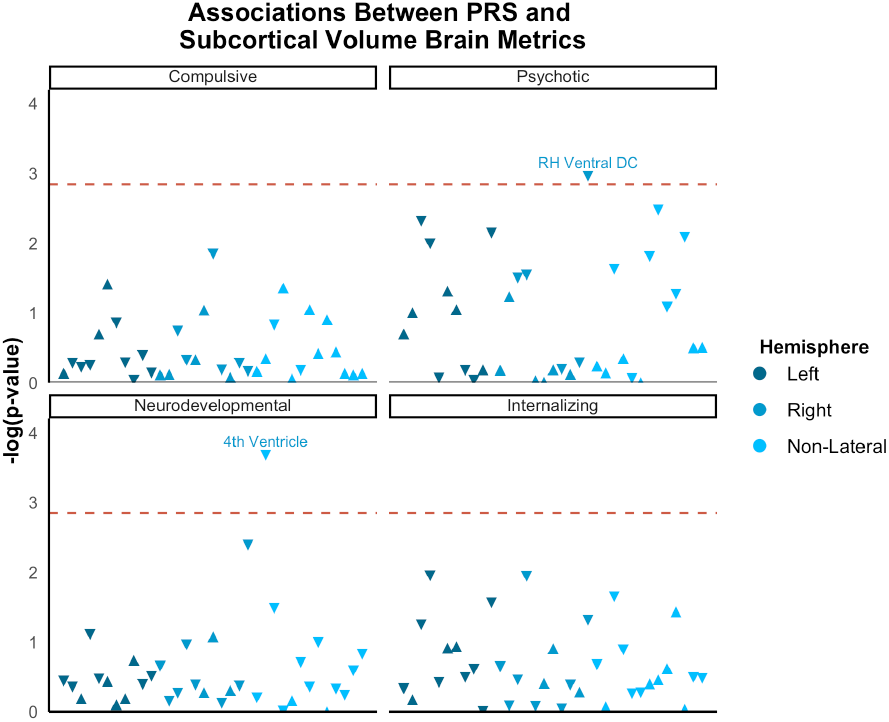
Associations between PRS and Subcortical Volume. Associations between each PRS and subcortical gray matter volume. The x-axes represent phenotypes, and the y-axis represent the -log_10_ of the uncorrected *p* values of associations between each PRS and each phenotype. Each dot represents one phenotype, with the colors indicating laterality and triangle direction indicating directionality of effect. The dashed lines indicate the threshold to survive Bonferroni correction. LH = left hemisphere; RH = right hemisphere; DC = diencephalon. Full results can be found in **Supplemental Table S17**.

**Extended Data Figure 9.**
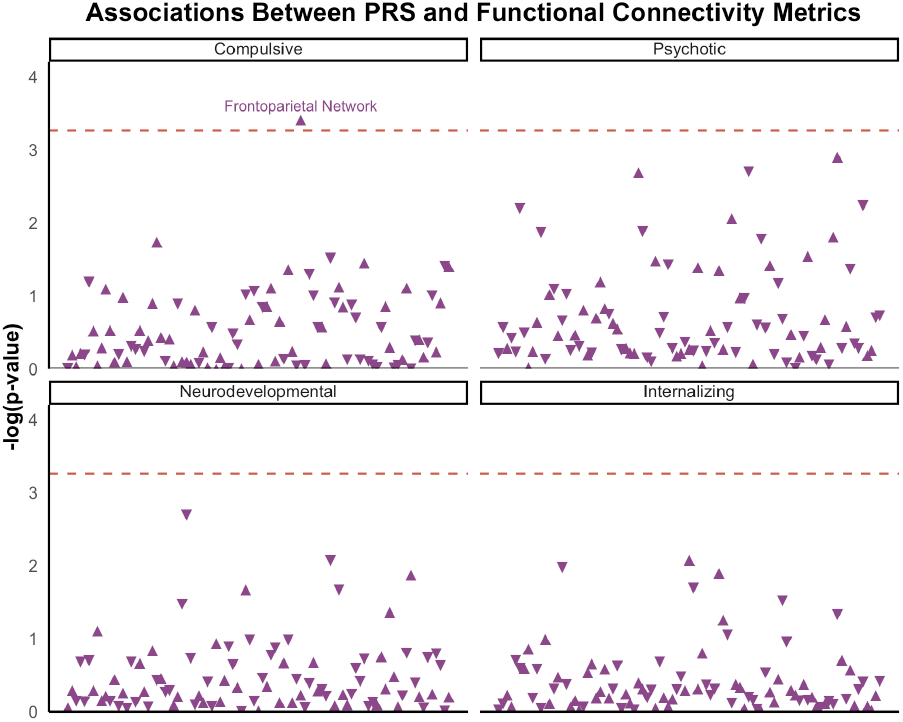
Associations between PRS and Functional Connectivity Metrics. Associations between each PRS and resting state functional connectivity metrics. The x-axes represent phenotypes, and the y-axis represent the -log_10_ of the uncorrected *p* values of associations between each PRS and each phenotype. Each dot represents one phenotype, with triangle direction indicating directionality of effect. The dashed lines indicate the threshold to survive Bonferroni correction. Full results can be found in **Supplemental Table S18**.

**Extended Data Figure 10.**
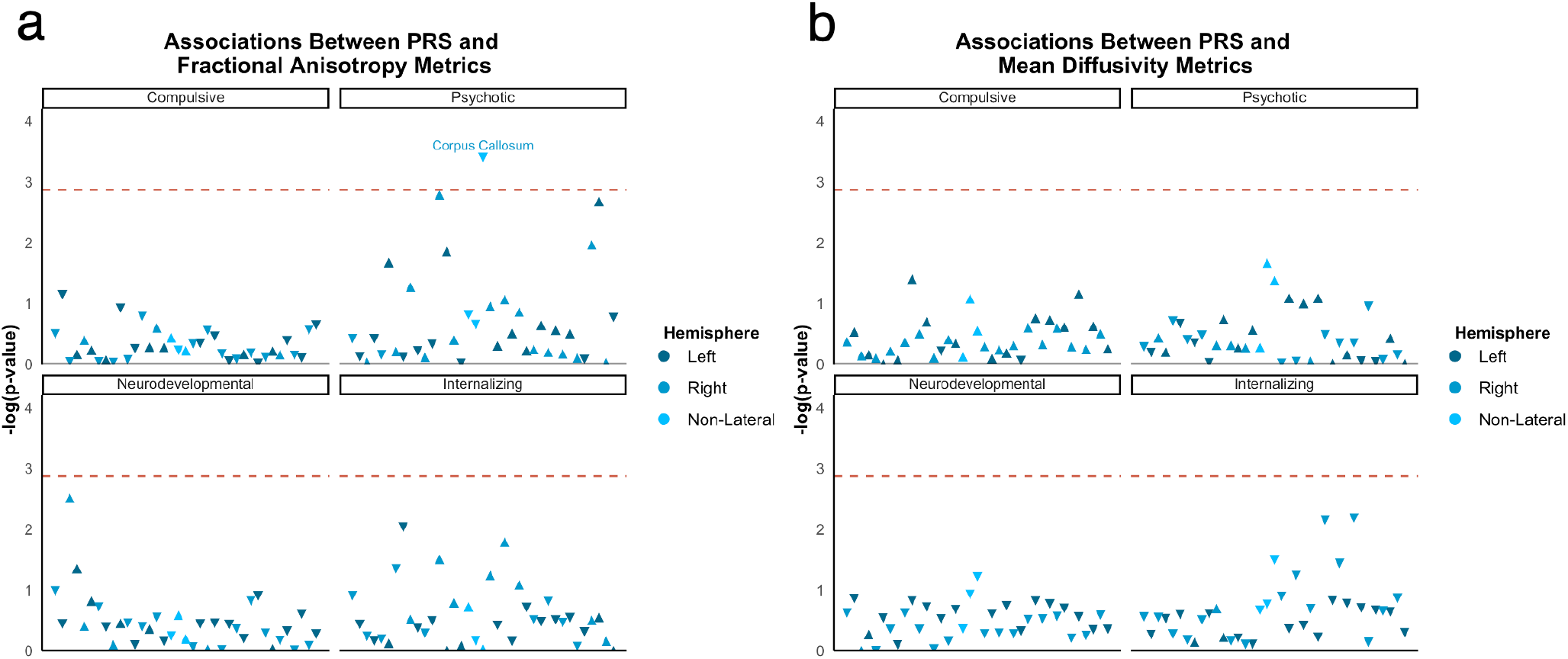
Associations between PRS and Diffusion Tensor Imaging Metrics. Associations between each PRS and diffusion tensor imaging metrics. **a)** Fractional Anisotropy. **b)** Mean Diffusivity. The x-axes represent phenotypes, and the y-axis represent the -log_10_ of the uncorrected *p* values of associations between each PRS and each phenotype. Each dot represents one phenotype, with the colors indicating laterality and triangle direction indicating directionality of effect. The dashed lines indicate the threshold to survive Bonferroni correction. Full results can be found in **Supplemental Tables S19-S20**.

