## Supplementary material for "Phenome-wide Investigation of Behavioral, Environmental, and Neural Associations with Cross-Disorder Genetic Liability in Youth of European Ancestry": Methods

**Genomic Structural Equation Modeling**

***GWAS Summary Statistics.*** Summary statistics from the largest available discovery GWAS were used for each of the 11 phenotypes of psychiatric disorder: Attention-Deficit/Hyperactivity Disorder (ADHD), Anorexia Nervosa (AN), Autism Spectrum Disorder (ASD), Bipolar Disorder (BIP), Generalized Anxiety Disorder (GAD), Major Depressive Disorder (MDD), Obsessive-Compulsive Disorder (OCD), Problematic Alcohol Use (PAU), Post-Traumatic Stress Disorder (PTSD), Schizophrenia (SCZ), and Tourette’s Syndrome (TS). Sample sizes and SNP heritability estimates are summarized in **Supplemental Table S6.**

For the current cross-trait GWAS, only SNPs present in all 11 GWAS were analyzed (i.e., SNPs that passed QC thresholds at all levels, resulting in 5,482,644 SNPs. LD scores were estimated from the European sample of 1000 Genomes, and analyses were restricted to HapMap3 SNPs, which tend to have higher imputation and heritability estimate accuracies. Effective Ns were estimated for each GWAS. For continuous traits (i.e., GAD and PAU), the given Ns were used. For case/control GWAS, the equation N_effective_ = 4v(1-v)n was used for each contributing cohort, were v = the proportion of cases and n = the total sample size; effective Ns for each contributing cohort were summed to provide the effective N for the GWAS.

*Anorexia Nervosa:* Summary statistics for AN were derived from a meta-analysis of GWAS of lifetime diagnosis of AN established via hospital or register records, structured clinical interviews, or online questionnaires based on DSM-III-R, DSM-IV, ICD-8, ICD-9, or ICD-10 criteria, or, as in the UK Biobank, via self-reported diagnosis^1^. GWAS were conducted separately on a total of 33 datasets (N = 16,992 cases; 55,525 controls) and meta-analyzed with the Ricopili pipeline inverse-variance weighted fixed-effect model. SNP-heritability for AN was estimated to be 0.11-0.17.

*Attention-Deficit/Hyperactivity Disorder:* Summary statistics for ADHD came from a meta-analysis of case/control GWAS in 11 PGC cohorts and in the population-based iPSYCH sample^2^. In iPSYCH, cases were diagnosed by psychiatrists according to ICD10 (F90.0). In the 11 PGC cohorts, ADHD was identified via multiple methods, including semi-structured diagnostic interviews and parent report. GWAS were conducted using logistic regression with imputed additive genotype doses in each sample, and GWAS were meta-analyzed using an inverse-variance weighted fixed effects model. The meta-analysis of individuals of European ancestry consisted of 19,099 cases and 34,194 controls, for an N_effective_ = 49,735. SNP-heritability was 0.216.

*Autism Spectrum Disorder:* ASD summary statistics came from a GWAS meta-analysis of 18,381 cases and 27,969 controls in the iPSYCH and PGC cohorts^3^. Cases in iPSYCH were identified from the Danish Psychiatric Central Research Register as having been diagnosed with ASD by a psychiatrist according to ICD10 (including childhood autism, atypical autism, Asperger’s syndrome, other pervasive developmental disorders, and pervasive developmental disorder, unspecified). Cases in the five PGC cohorts had ASD diagnoses identified from standard research tools or expert clinical consensus. Variants were filtered to MAF ≥ 0.01 and INFO score ≥ 0.70. The meta-analysis was conducted in METAL with an inverse-variance-weighted fixed-effect model. The N_effective_ is 43,777, and SNP-heritability was estimated to be 0.118.

*Bipolar Disorder:* Summary statistics for BIP were derived from a GWAS meta-analysis of 41,917 cases with bipolar disorder and 371,549 controls across 57 cohorts, for an N_effective_ of

101,962 individuals^4^. Across 52 cohorts, cases were identified according to international consensus criteria for a lifetime BIP diagnosis established with structured diagnostic assessments, medical record review, or clinician-administered checklists. For five cohorts (i.e., iPSYCH, deCODE, Estonian Biobank, HUNT, and UK Biobank), cases were determined via ICD codes or self-report. See^4^ for descriptions of QC procedures in each of the cohorts. GWAS within each cohort were conducted using additive logistic regression models and meta-analyzed in METAL with an inverse-variance-weighted fixed-effects model. SNP-heritability was estimated to be .156 and .186, assuming population prevalence of 2% and 1%, respectively.

*Generalized Anxiety Disorder:* GAD summary statistics came from a GWAS meta-analysis of Generalized Anxiety Disorder 2-item scale (GAD-2) in 175,163 individuals in the Million Veterans Program^5^. SNPs with MAF < 0.001 or INFO < 0.3 were excluded from analysis. The GWAS was conducted with linear regression in two tranches within the European ancestry subsample and meta-analyzed with inverse variance weighting in METAL. SNP-heritability was found to be 0.0558.

*Major Depressive Disorder:* Summary statistics for MDD came from a GWAS meta-analysis of 170,756 cases with MDD and 329,443 controls (N_effective_ = 449,149) in the UK Biobank and PGC^6^. Cases in the UK Biobank were defined according to self-reported treatment-seeking for “nerves, anxiety, tension or depression,” and in the PGC according to structured diagnostic interviews. Variants with MAF < 0.005 or INFO < 0.1 in the UK Biobank, and with MAF < 0.005 or INFO < 0.6 in the PGC, were excluded. METAL was used to conduct an inverse variance-weighted meta-analysis. SNP-heritability was found to be 0.089.

*Obsessive Compulsive Disorder:* OCD summary statistics were taken from a GWAS meta-analysis of OCD cases and controls from IOCDF-GC and OCGAS, totaling 2,688 cases and 7,037 controls (N_effective_ = 7,281)^7^. All cases met criteria for OCD according to the DSM-IV diagnostic criteria. SNPs with MAF < 0.01 or INFO < 0.6 were excluded Separate association analyses were conducted for each case-control subpopulation (e.g., Ashkenazi Jewish) and meta-analyzed using METAL with the inverse variance method. SNP-heritability was estimated to be 0.25.

*Posttraumatic Stress Disorder:* Summary statistics for PTSD came from a GWAS meta-analysis of 23,212 lifetime or current PTSD cases (according to versions of the DSM) and 151,447 controls (majority trauma-exposed) of European ancestry that were part of the PGC-PTSD Freeze 2 dataset (N_effective_ = 70,332)^8^. SNPs were filtered to exclude MAF < 0.10 and INFO < 0.6, and meta-analyses within and across ancestral groups were performed with METAL using inverse variance weighted fixed effects. SNP-heritability was 0.05.

*Problematic Alcohol Use:* PAU summary statistics were derived from a GWAS meta-analysis of alcohol use disorder and problematic drinking in 435,563 individuals of European ancestry (N_effective_ = 300,789)^9^. Contributing GWAS included two GWAS of ICD-defined AUD in the Million Veterans Program, a GWAS of DSM-IV alcohol dependence in the PGC, and a GWAS of the AUDIT-P in the UK Biobank. Variants with MAF ≤ 0.001 or INFO ≤ 0.7 in MVP, MAF ≤ 0.01 or INFO ≤ 0.8 in the PGC, and MAF < 0.001 or INFO < 0.7 in the UKB were excluded. The sample size-weighted method in METAL was used to meta-analyze the summary statistics. SNP-heritability for PAU was estimated to be 0.068.

*Schizophrenia:* The most recent SCZ GWAS meta-analysis of 67,390 schizophrenia/schizoaffective disorder cases (80% European ancestry, 20% East Asian ancestry) and 94,015 controls (N_effective_ = 157,013) from the PGC was used^10^. Individual cohort association analyses were conducted using additive logistic regression models with PLINK and meta-analyzed with a standard error inverse-weighted fixed effects model. SNP-based heritability for SCZ was found to be 0.24.

*Tourette Syndrome:* TS summary statistics came from a GWAS meta-analysis of 4,819 Tourette syndrome cases and 9,488 controls (N_effective_ = 12,140) from four European ancestry datasets^11^. Cases were identified by DSM-IV-TR criteria and clinician observation, web-based phenotypic assessments, specialty clinics, or DSM-5 criteria for Tourette syndrome or chronic motor or vocal tic disorder. The GWAS meta-analysis was conducted using the inverse-variance method in METAL. SNP-heritability was estimated to be 0.21.

***Confirmatory Factor Analysis.*** The GenomicSEM^12^ package in R was used to run a multivariate GWAS of 11 psychiatric phenotypes (See **Supplemental** **Table S6** for descriptions of the contributing GWAS). First, the publicly available summary statistics were formatted for preprocessing. Second, using LD weights for European ancestry from 1000 genomes and the LD score regression function of GenomicSEM were used to calculate the genetic covariance matrix. As depicted in **Extended Data Fig. 1,** genetic correlations among the psychopathology phenotypes ranged from -0.17 between ADHD and OCD to 0.95 between MDD and GAD, with most pairs of phenotypes demonstrating significant genetic correlations. Confirmatory factor analyses based on models identified in Grotzinger et al. were fit and assessed according to values on the comparative fit index (CFI; values > 0.95 indicate good fit) and standardized root mean square residual (SRMR; values < 0.08 indicate good fit). The original correlated factors model fit the data well (CFI = 0.952, SRMR = 0.0783). Because the loading of MDD onto the Neurodevelopmental factor was negative (-0.23), it was removed. Model fit remained excellent (CFI = 0.966, SRMR = 0.080, chisq (34) = 216). Finally, given that the covariance between the Compulsive and Neurodevelopmental factors was zero, the covariance was explicitly constrained to zero, and model fit remained excellent (CFI = 0.966, SRMR = 0.080, chisq (35) = 216) The output of this model is shown in **Figure 2.** The hierarchical model presented in Groztinger et al. (2022) was also tested, showing generally poorer fit (CFI = 0.947, SRMR = 0.087, chisq (35) = 381). See **Supplemental Tables S24** and **S25** for a description of model fits and factor loadings.

***Multivariate GWAS.*** Next, a multivariate GWAS was conducted based on this final model with the userGWAS() function in GenomicSEM, using the 5,482,644 SNPs available across the 11 phenotypes. The genomic SEM model specified above was expanded to include SNP effects that occur at the level of the latent genetic factors. Indicators were constrained to have variances greater than 0.001 to aid in model convergence. Then, one multivariate GWAS per latent factor was conducted in which the SNP effects were specified to operate through the genetic indicators and not the common factors. The resulting chi-square values were then subtracted from the common pathways model chi-square values, and p-values were calculated using the degrees of freedom difference between the models. Significant Q-SNPs have effects that are not mediated by the latent genetic factors (i.e., effects on latent factors are driven by heterogeneous effects across indicators) and were removed from the summary statistics before polygenic risk scoring, along with those SNPs in linkage disequilibrium (r^2^ threshold of 0.1, 500-kb window; Grotzinger et al.^12^). These summary statistics were uploaded to FUMA, and 11, 224, 14, and 78 independent genomic risk loci were identified for the Compulsive, Psychotic, Neurodevelopmental, and Internalizing factors, respectively (see **Supplemental Tables S7-S10** for more information). The total effective sample sizes for the factors were: Compulsive (n = 39,364), Psychotic (n = 160,932), Neurodevelopmental (n = 37,665), and Internalizing (n = 432,638). See **Extended Data Figs. 2-3** for GWAS Manhattan plots and **Supplemental Note** for further characterization of GWAS results.

**Polygenic Risk Scores**

***Genotyping, Quality Control, and Imputation.*** Saliva samples were genotyped on the Smokescreen array^13^ by the Rutgers University Cell and DNA Repository (now incorporated with other companies as Sampled; https://sampled.com/). The Rapid Imputation and COmputational PIpeLIne for Genome-Wide Association Studies (RICOPILI^14^) was used to perform quality control (QC) on the 11,099 individuals with available ABCD Study phase 3.0 genotypic data, using RICOPILI’s default parameters. The 10,585 individuals who passed QC checks were matched to broad self-reported racial groups using the ABCD Study parent survey. Of the 6,787 parents/caregivers indicating that their child’s race was only “white,” 5,561 of those individuals did not endorse any Hispanic ethnicity/origin. After performing a second round of QC on these sub-samples, 5,556 non-Hispanic White individuals were retained in the analyses. Principal component analysis (PCA) in RICOPILI was used to confirm the genetic ancestry of these individuals by mapping onto the 1000 Genomes reference panel, resulting in a PCA-selected European-ancestry subset. The European ancestry subset was then imputed to the TOPMed imputation reference panel^15^. Imputation dosages were converted to best-guess hard-called genotypes, and only SNPs with Rsq > 0.8 and MAF > 0.01 were kept for PRS analyses.

***Polygenic Risk Scoring.*** PRS-CS^16^ and PLINK v2.0 ^17^ were used to compute polygenic risk scores (PRS) for the four higher order latent genetic factors. PRS-CS is a Bayesian polygenic prediction method that applies a continuous shrinkage prior to SNP effect estimates and infer posterior SNP weights. Here, PRS-CS was used to adjust the weights for the 836,306–839,299 overlapping SNPs, with 1000Genomes used as reference panel for LD. The PLINK –score command was used to calculate PRS for each individual by summing all variants weighted by the inferred posterior effect size.

**Phenome-wide Association Study (PheWAS)**

***Non-Imaging Phenotypes.*** Phenotypes for the PRS-PheWAS were selected from a comprehensive sample of items measured in the ongoing longitudinal Adolescent Brain Cognitive Development^SM^ (ABCD) Study, which follows 11,875 children recruited at baseline (ages 8.9-11 years) from 21 research sites across the United States. Parents/caregivers provided written informed consent, and children verbal assent, to a research protocol approved by the institutional review board at each of 21 data collection sites across the United States (<https://abcdstudy.org/sites/abcd-sites.html>). For the present analyses, data were drawn from data release 3.0 for baseline assessment phenotypes and from data release 4.0 for two-year follow-up (FU2) phenotypes.

Rigorous quality control procedures were implemented to ensure sufficient endorsement and relevance of each phenotype. See **Supplemental Tables S1**-**S5** for notes on specific measures and item retention and recoding. A total of 6,407 and 10,129 non-imaging variables were considered at baseline and FU2, respectively, spanning the domains of substance use, mental health, physical health, neurocognition, culture and environment, demographics, and screen time. Phenotypes were examined for relevance (e.g., item redundancy, such as raw and t-scores; administrative items, such as assessment device and items indexing the number of questions answered), and irrelevant items were removed (1,624 removed at baseline, 5,751 at FU2). Following previous studies^18^, variables with fewer than 100 data points or fewer than 100 endorsements of minority categories of categorical variables were then removed, leaving 1,265 and 1,687 items at baseline and FU2, respectively. Several multi-category items were dummy coded, bringing the total items to 1,269 and 1,694, respectively, of which 799 phenotypes were assessed at both waves. These phenotypes consisted of proximal (i.e., diagnoses and symptoms of constituent disorders and related traits, such as disinhibition and executive function) and more distal (i.e., substance use and access, environmental exposures and experiences, physical health, electronic media use, other cognitive functioning). To enhance accuracy, this task was split among multiple investigators (SEP, NRK, ISH, AJG), and each checked over the others’ work and provided consensus that retained variables were relevant.

Below, the measures included in each domain are briefly summarized.

*Cognition.* At both waves, all available uncorrected summary scores from the NIH Toolbox Cognitive Battery were included, as were summary scores for the Little Man Task and Rey Auditory Verbal Learning Task. Matrix Reasoning and the Cash Choice Task measures were additionally available at the baseline assessment.

*Screen Time.* Caregiver- and youth-reported screen time metrics were included at both waves, with additional items available at FU2. The questionnaires ask about time spent on screens for various types of media, separated for weekdays and weekend days, as well as the frequency of certain screen media activities (e.g., mature-rated video games). At follow-up, new items assess effects of screen time (e.g., causing arguments, losing track of time spent on phone, difficulty discontinuing use of screen media) as well as social media-specific behaviors and effects.

*Demographics.* Caregiver-reported demographic variables, such as income, education, employment, financial difficulties, among others, were included at each wave, with financial difficulty phenotypes at FU2 coming from a longitudinal assessment.

*Substance Use and Related Phenotypes.* At each wave, youth self-reported substance familiarity and use patterns, as well as intention to use, subjective response to various substances, and peer use patterns are assessed. Youth-reported peer tolerance of use and perceived harm of using particular substances, as well as consequences of use, are additionally included at FU2. Further, objective measures of use, including breathalyzer, nicalert, hair tests, and other toxicology tests are available for all or a subset of participants at each wave. Finally, measures of parental rules about substance use and community risk and protective factors are also assessed.

*Culture/Environment.* At both waves, environmental variables included the youth- and caregiver-reported family environment scale, neighborhood safety and environment, and youth prosocial behavior; youth-reported parental monitoring, and school risk and protective factors; and neighborhood environment measures as assessed on the residential history derived scores. At baseline, youth also completed the children’s report of parental behavior inventory. Additional measures at FU2 included the youth discrimination measure, youth peer behavior and peer network health scales, youth- and parent-reported school attendance and grades, parent-reported community cohesion, youth- and parent-reported life events, a measure of cyber bullying, peer experiences, and the parent occupation survey.

*Physical Health.* Physical health measures at both assessment time points consisted of medical history (e.g., broken bones, ER visits), medication inventory, caregiver-reported sleep disturbances, youth- and caregiver-reported pubertal development, traumatic brain injury information, sports and activities involvement, and anthropometrics (i.e., height, weight, waist circumference). Baseline measures also included the developmental history questionnaire, which assesses prenatal, pregnancy, and birth events and exposures, and pubertal hormones (i.e., testosterone, estradiol, and DHEA. At follow-up, additional measures included blood analysis, blood pressure, chronotype, pain, reported and measured physical activity, and dietary intake.

*Family Mental Health.* Family mental health consists of items reflecting family history and caregiver symptoms and behaviors. Family history was only assessed at baseline, both waves included the Adult Self Report, and FU2 added the Adult Behavior Checklist.

*Child Mental Health.* This category contains the most phenotypes at both baseline and FU2. At both waves, measures included the CBCL, Brief Problem Monitor-Teacher Report, UPPS-P, Prodromal Psychosis Scale, the General Behavior Inventory-Mania, Resilience, and the Bis/Bas System Scales. The KSADS-5 was also administered at both waves, with slightly different items released at FU2 (i.e., items regarding frequency and impairment used to determine if symptoms met diagnostic threshold). At FU2, the youth Brief Problem Monitor, 7-Up Mania items, NIH Toolbox Positive Affect items, the early adolescent temperament questionnaire, and the short social responsiveness scale were added.

***Imaging Phenotypes.*** Imaging data are available for 11,556 out of 11,875 participants. A detailed description of the imaging acquisition procedures and processing and analysis methodology in ABCD can be found elsewhere^19,20^. Briefly, 1mm isotropic T1-weighted structural images were obtained via 3T MRI scanners (Siemens, Phillips, and GE) using either a 32- or 64-channel head-and-neck coil and completed T1-weighted and T2-weighted structural scans (1mm isotropic). MRI scan protocols were harmonized across the three MRI vendor platforms to minimize variability. Real-time motion detection and correction was implemented to mitigate the influence of head motion. Hagler et al. (2019)^20^ provides a comprehensive description of the quality-control measures conducted on the processed imaging data.

For structural MRI (sMRI) metrics, structural neuroimaging processing was completed using FreeSurfer version 5.3.0 through standardized processing pipelines^20^. Cortical reconstruction and volumetric segmentation was performed by the **ABCD Study^®^** Data Acquisition and Integration Core using the FreeSurfer image analysis suite (<http://surfer.nmr.mgh.harvard.edu/>). This pre-processing includes removal of non-brain tissue using a hybrid watershed/surface deformation procedure^21^, automated Talairach transformation, segmentation of the subcortical white matter and deep gray matter volumetric structures, intensity normalization, tessellation of the gray/white matter boundary, automated topology correction, and surface deformation following intensity gradients^20^. Images were registered to the Desikan atlas, which was based on individual cortical folding patterns to match cortical geometry across subjects. The cerebral cortex was parcellated into 34 regions per hemisphere based on the gyral and sulcal structure. For the imaging analyses, cortical gray matter volume, thickness, and surface area, and total cerebral white matter volume aligned to the Desikan atlas were extracted from ABCD data release 3.0. Fifteen global metrics were analyzed first along with two diffusion MRI (dMRI) variables and included total and bilateral cortical volume and surface area, mean cortical thickness, total subcortical gray matter volume, intracranial and whole brain volume, and supratentorial volume, as well as bilateral cerebral white matter volume. An additional 239 regional sMRI metrics (i.e., 34 cortical regions*2 hemispheres*3 metrics (i.e., volume, surface area, thickness) + 35 subcortical regions (i.e., accumbens, thalamus, caudate, putamen, pallidum, hippocampus, amygdala, ventral diencephalon, brainstem, cerebellum, ventricles, cerebrospinal fluid, corpus callosum segments) were extracted for potential follow-up. Only sMRI data that passed these QC tests (n=5,310) were retained.

For diffusion MRI (dMRI), we examined mean diffusivity (MD) and fractional anisotropy (FA) metrics. dMRI metrics were calculated with a linear estimation approach with log-transformed

diffusion-weighted signals^22^. To create MD and FA metrics, first, tensor matrices were diagonalized using singular value decomposition, resulting in three eigenvectors and three corresponding eigenvalues. FA is derived from the eigenvalues^22^. MD is derived from the mean of the eigenvalues. Major white matter tracts are labelled using AtlasTrack, a probabilistic atlas-based method for automated segmentation of white matter fiber tracts^23^. For dMRI, we examined 2 summary metrics (total fractional anisotropy and mean diffusivity for all fibers) and 37 regional metrics each for FA and MD. Only dMRI data that passed QC tests (n=4,924) were retained.

Resting state functional connective (rs-fMRI) metrics were also extracted for analyses. For rs-fMRI data collection, participants completed four 5-minute resting-state BOLD scans, with their eyes open and fixated on a crosshair. Resting state images were acquired in the axial plane using an EPI sequence. Other resting-state image parameters varied by 3T scanner and have been previously detailed (<https://abcdstudy.org/images/Protocol_Imaging_Sequences.pdf>)^19^.

A data analysis pipeline, using the Multi-Model Pressing Stress software package, was created to analyze RSFC data^20,24^. In terms of motion correction, motion regression included six parameters plus their derivatives and squares. Only frames with FD<0.3mm were included in the regression. Additionally, all models examining rs-fMRI metrics included average motion. Pair-wise correlations were examined for ROIs within functionally-defined parcellations (i.e., Gordon networks)^25^. The Fisher Z-transform of the correlation values were examined within each of the 13 Gordon networks across 78 additional between-network rs-fMRI metrics). Only rs-fMRI data that passed QC tests (n=5,255) were retained.

***PheWAS Analyses.*** Of 11,875 children (mean ± SD age = 9.91±0.62 years; 47.85% female; 52.1% white) who completed the baseline assessment of the ongoing longitudinal Adolescent Brain Cognitive Development^SM^ (ABCD) Study, 5,556 participants of genomically-confirmed European ancestry were included in the present analyses (mean ± SD age = 9.93 ±0.63 years; 47.0% female). At two-year follow-up (FU2; release 4.0), data came from 5,048 participants (mean ± SD age = 12.02±0.66 years; 46.93% girls; see **Table 1** for a summary of sample characteristics). Mixed-effects regression models were used to test associations between each of the four latent factor PRS and 1,269 and 1,694 phenotypes at baseline and two-year follow-up. Participants of non-European genomic ancestry were excluded (n=6,319) from analyses due to the lack of a well powered ancestry-specific discovery GWAS of the majority of psychiatric phenotypes included here in other ancestries, the relatively uninformative and low predictive utility of PRS when applied across ancestries^26^. All continuous variables were standardized and scaled prior to the analyses. The lme4 package^27^ in R was used to run linear (for continuous outcomes) and generalized linear (for dichotomous outcomes) mixed effects models, with random intercepts included for family and study site. For baseline analyses, covariates included age at assessment, sex, and the first 10 ancestral principal components; follow-up analyses additionally included a variable indicating whether the assessment had been in-person, remote, or hybrid given the COVID-19 pandemic. Models that did not converge due to lack of variability in covariates or random effects were modified to exclude problematic parameters (e.g., covariate of sex was removed from pubertal development models analyzing boys and girls separately). Bonferroni correction was used to correct for the number of tests (i.e., 1,269 and 1,694), separately within each PRS. False discovery rate (FDR)-corrected p-values are also presented, as the Bonferroni approach may be overly conservative given correlations among phenotypes.

Linear mixed-effects models were also used to test associations between each of the PRS and global imaging metrics, which were also standardized and scaled. In addition to the covariates noted above, MRI scanner type and motion (for rs-fMRI and dMRI analyses) were included. Bonferroni correction was applied to all p-values within each modality (i.e., global, cortical volume, surface area, thickness, subcortical volume, rs-fMRI, dMRI FA, dMRI MD) within each PRS model. In models examining regional cortical volume, total cortical volume was included as a covariate. Total surface area was included in regional surface area models, and mean thickness was added in regional thickness models. Total fractional anisotropy across all fibers was included as a covariate in all dMRI models.

***Indirect Effects Analyses.*** In order to explore whether baseline imaging metrics indirectly linked PRS to two-year follow-up phenotypes, associations between baseline imaging metrics and two-year follow-up phenotypes were first tested. Only metrics and phenotypes that were significantly associated with the same PRS were included, and linear mixed effects models covaried for baseline age, sex, MRI scanner type, COVID visit type, and nested by family and site ID. For imaging metrics showing significant associations with both, the lavaan^28^ and lavaan.survey^29^ R packages were used to test whether they indirectly linked PRS to follow-up phenotypes. We fit a path analysis model that included the direct effect of PRS on follow-up phenotype as well as the indirect effect of PRS on follow-up phenotype via imaging metric, using Maximum Likelihood for continuous phenotypes and the weighted least squares mean and variance estimator (WLSMV) for dichotomous phenotypes. Endogenous dichotomous phenotypes were specified to be ordered (i.e., “ordered = TRUE” in lavaan). On the first path from PRS to imaging phenotypes, covariates included the first 10 ancestral PCs, baseline age, sex, and scanner type. On the path from PRS to follow-up phenotype, controlling for imaging metric, covariates included the first 10 ancestral PCs, follow-up age, sex, scanner type, and COVID visit type. The svdesign() function in the survey R package^30^ was used to specify groups for family and site ID, and the lavaan.survey() function used the output from the original path analysis to refit the model nesting by the specified groups. Bonferroni correction was used to correct for the number of indirect effects models conducted within each imaging metric.
