## Supplemental Note for "Phenome-wide Investigation of Behavioral, Environmental, and Neural Associations with Cross-Disorder Genetic Liability in Youth of European Ancestry"

**Multivariate GWAS Results**

Upon uploading the GWAS summary statistics for each individual factor to FUMA, we identified a total of 38 significant independent lead SNPs in 37 independent Q_SNP_ loci for the Compulsive factor. Grotzinger et al. identified two significantly heterogeneous loci on chromosomes four and six, one of which was also identified in the current study. The heterogeneous Q_SNPs_ identified were removed from the Compulsive factor summary statistics used in follow-up analyses. We identified 12 independent lead SNPs in 11 independent genomic risk loci for the Compulsive factor. Grotzinger et al. Identified 1 independent hit for their Compulsive factor, and this locus was identified in our study. Additionally, the current study identified 10 novel hits associated with our Compulsive factor, but not the Compulsive factor of the previous study.

A total of 38 significant independent lead SNPs in 37 independent Q_SNP_ loci for the Psychotic factor were identified. Grotzinger et al. identified six significantly heterogeneous loci on chromosomes two, three, four, and 12, two of which were also identified in the current study. The heterogeneous QSNPs identified were removed from the Psychotic factor summary statistics used in follow-up analyses. We identified 285 independent lead SNPs in 224 independent genomic risk loci for the Psychotic factor. Grotzinger et al. Identified 108 independent hits for their Psychotic factor, of which 90 were identified in our study. Here, we identified 134 novel hits associated with our Psychotic factor, but not the Psychotic factor of the previous study. That the present study identified more hits for the Psychotic factor than the previous Grotzinger et al. GWAS may be attributable to our selection of more recent and better-powered GWAS for Schizophrenia (i.e., 67,393 vs 53,386 cases) and Bipolar Disorder (i.e., 41,917 vs 20,352 cases).

We identified a total of 38 significant independent lead SNPs in 37 independent Q_SNP_ loci for the Neurodevelopmental factor. Grotzinger et al. identified seven significantly heterogeneous loci on chromosomes two, three, four, and six, and eight, one of which was also identified in the current study. The heterogeneous QSNPs identified were removed from the Neurodevelopmental factor summary statistics used in follow-up analyses. We identified 15 independent lead SNPs in 14 independent genomic risk loci for the Neurodevelopmental factor. Grotzinger et al. Identified 9 independent hits for their Neurodevelopmental factor, of which 5 were identified in our study. Additionally, the current study identified 9 novel hits associated with our Neurodevelopmental factor, but not the Neurodevelopmental factor of the previous study.

We identified 32 significant independent lead SNPs in 29 independent Q_SNP_ loci for the Internal factor. Grotzinger et al. identified three significantly heterogeneous loci on chromosome four, all three of which were also identified in the current study. The heterogeneous Q_SNPs_ identified were removed from the Internal factor summary statistics used in follow-up analyses. We identified 95 independent lead SNPs in 78 independent genomic risk loci for the Internal factor. Grotzinger et al. Identified 44 independent hits for their internalizing factor, and 23 of these loci were identified in the current study. We identified 55 novel hits associated with our Internal factor, but not the Internalizing factor of the previous study. That the present study identified more hits for the Internalizing factor than the previous Grotzinger et al. GWAS may be attributable to our selection of more recent and better-powered GWAS for Anxiety (i.e., N=175,163 vs 100,876) and PTSD (i.e., 23,212 vs 12,255 cases).
